# Recurrent Group-switch Interactions in Heterogeneous Population Epidemic Modelling

**DOI:** 10.1101/2025.11.13.25340210

**Authors:** Michael Smah, Anna Seale, Kat Rock

## Abstract

Unexpected epidemic outcomes that show discrepancies between predicted and actual epidemics and the impacts of interventions are often attributed to factors such as biased data collection or pathogen evolution, while less attention is given to the realism of the epidemic models that inform public health decisions. Many models guiding policy still rely on simplified assumptions of homogeneous mixing, usually combining the effects of mobility and differing contact strengths within/between different sociodemographic groups at home and at work/school into a single transmission parameter, thereby overlooking the heterogeneity and structure of real human contact networks. Even in age-structured models, people within the same age group are considered to mix equally; moreover, differences in inter-group interaction that exist at home and work/school are often ignored. Recent studies have improved the realism of epidemic modelling by incorporating the concept of semi-random mixing and explicitly representing household and non-household interactions linked by daily mobility. Building on these developments, this study introduces a recurrent group-switch epidemic model that captures how individuals transition between socio-demographic groups at home and work or school, incorporating sociodemographic structure and semi-random mixing within a computationally efficient equation-based framework. Analytical derivations yield new formulations for the force of infection and group-specific metrics, including source, sink, and source-to-sink reproduction numbers. Model simulations using UK age-structured contact data underscore how, for COVID-19-like infections, individuals aged 6–24 years act as key drivers of transmission, while older adults serve primarily as infection recipients rather than infectors (sinks) with higher hospitalisation risks. Modelling non-pharmaceutical interventions shows that reducing inter-household/cluster connectivity among younger populations may substantially reduce transmission, whereas mobility restrictions alone can produce counterintuitive increases in epidemic size. By explicitly linking recurrent social behaviour, heterogeneity, and mobility, this model framework improves the realism of epidemic models and provides deeper insight into group-specific transmission dynamics, which could be used to guide more targeted and effective public health interventions.

## 1 Background

The classical compartmental epidemiological models, such as the Susceptible-Infected-Recovered (SIR) model and its variations, have contributed to understanding and predicting the course of epidemics and the design and analysis of interventions. To enhance the predictive capabilities of epidemic models, the modelling formulation should reflect the realism of human interaction and heterogeneity [1–3]. When modelling diseases that spread unevenly through populations or when assessing targeted interventions (like prioritising vaccinations or limiting movement), it can be very important to include realistic heterogeneity, such as differences in contact rates, spatial mobility, demographic structure, and behavioural responses. Taking this kind of heterogeneity into account often makes model results easier to understand and more useful for policy by showing how the mechanisms behind observed epidemic dynamics work. Incorporating realistic aspects of epidemic processes and human interaction can complicate mathematical analysis and potentially raise the computational cost of numerical simulations [4]. Detailed or heterogeneous models may face the issue of over-parameterisation, where it becomes challenging or impossible to reliably estimate parameters based on available data. In situations requiring urgent public health decisions, simpler models that ensure quick computation and straightforward interpretation may be favoured over detailed ones that require more time for implementation and mathematical analysis.

It is a common practice to make simplifying assumptions; for example, the assumption that people are exposed to the risks of infection equally, regardless of their age, gender or any socio-economic differentials [5]. For the effectiveness of targeted interventions, it is important to acknowledge that, in any population, there is heterogeneity, which may include differences in age, socioeconomic status, location, occupation, and existing health conditions. This heterogeneity influences infectious pathogen spread and may be reflected in differences in infection rates as well as variations in disease severity and health outcomes. For example, during the COVID-19 pandemic, children were at a higher risk of infection, but their disease severity was typically mild, whilst older people had an increased risk of severe disease [6].

Risk-structured epidemic modelling [7–9] is often used to incorporate differing risk levels among individuals, reducing the generalisation of homogeneous risk in simple models. These risk-structured models classify people based on socio-demographic differences such as age, sex, and occupation, which can reflect their interactions and potential risk of infection. This method is used to help identify high-risk groups, which can inform public health interventions.

Although risk-structure epidemic models do not assume uniform random mixing between different population groups, they typically assume that within the same group, everyone is equally likely to meet with everyone else. This assumption overlooks the fact that even within the same population group, people only meet with a limited pool of potentially recurrent contacts (e.g., within households, work, and school). Another simplifying assumption typical of the risk-structure equation-based models is that the differing strengths of interaction within and between groups at home and when they go out to work/school are often ignored.

The semi-random mixing (SeRaMix) equation-based model (EBM) has been proposed in the literature [10] to relax the standard random mixing assumption. The study, although it did not incorporate a risk structure, captures one key reality about human interactions: people have a limited pool of individuals (*potentially-recurrent contacts*), for example, household members, school classmates, and office colleagues. People sharing the same cluster (household, office, or school) are typically daily potential contacts of each other. Thus, each person in a cluster of size *n* will have *n* − 1 connections within the cluster. In addition to this, each person will have *x* external connections with whom daily contact is likely. During each time step, each person chooses *c* ≤ *n* − 1 + *x* random contacts from their *n* − 1 + *x* connections, such that people do not necessarily have the same contacts every time and do not make random contacts in the population, as the pool of potential contacts is limited to one’s potentially recurrent contacts, *n* − 1 + *x*. In the context of risk-structured modelling, the potentially recurrent contacts of any individual may include a mix of people from different population groups, from which usual contacts may be randomly selected, but the strength of within/between contacts may differ with population groups.

Using the strength of the SeRaMix model [10], an explicit interaction of home and work/school connectivity was proposed [11] to model disease propagation in a multi-clustered population to introduce another reality of human interaction: human daily interactions are multi-phased. In this EBM formulation, a day starts with household interactions, followed by either daytime household interaction for household members that remain at home or daytime work/school interaction for those that move out of the household. All individuals then return home from work/school, and the pattern begins again. The rate of within- and between-group interaction may differ depending on whether interactions happen at home or work/school. For example, interactions between children and older people at home may be different from those at work/school. The pool of children from which a ‘typical’ child may choose their contacts when at home may be different from the pool at school. These realities have not been explored in the previous literature that we explored, and require attention with the view to improving EBMs for infectious disease modelling.

In this study, we introduce the concept of recurrent group-switch behaviour in epidemic modelling as described in the schematic diagram in Figure 1. The diagram depicts how households are made up of different groups (e.g., age groups, work groups, and other risk groups) that often disintegrate during work time, with each household member going to interact with a different group(s) before returning home, thereby switching between household groups and work groups every day. Thus, at each time step, each individual has more than a single preference for contacts if such a person can interact both within and outside the household. For example, the interaction of children with adults could be different depending on whether the interactions occur within the households where children interact with their guardians or outside the household, where children are more likely to interact with other children (e.g., at school). Similarly, the interaction between a banker and a teacher would depend on whether they interact at home, where contact quality (duration, proximity) is irrespective of occupation, or whether they interact outside the home, where they are more likely to interact with those in the same profession than with others. By incorporating recurrent group-switch behaviours, reflecting interactions closer to reality, this EBM captures how heterogeneous (group-dependent) mobility and interaction affect pathogen spread differently for different groups, enabling the analysis of the role of individual groups in the spread of infections.

**Figure 1.**
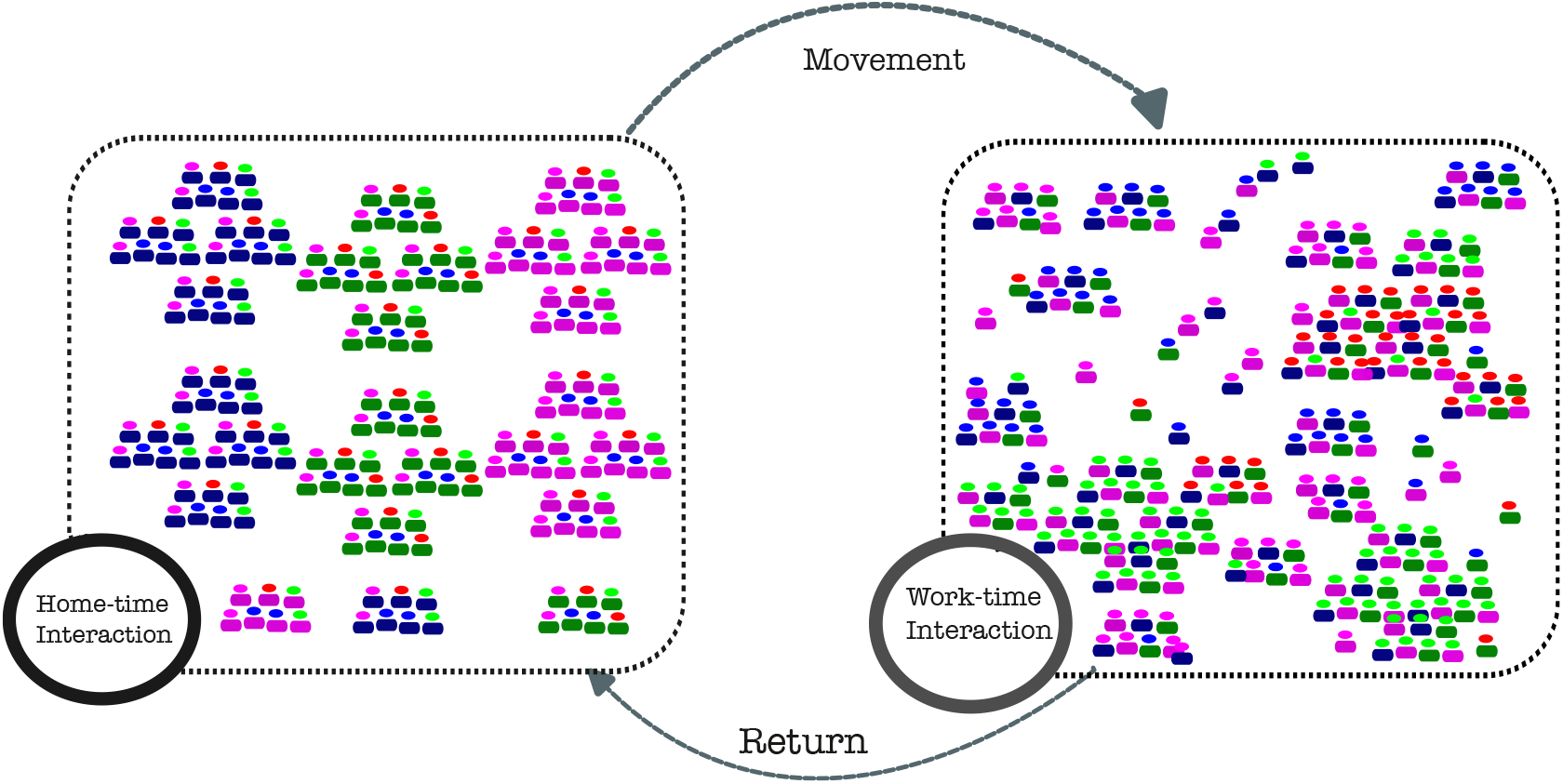
Schematic representation of the recurrent Group-switch model. The colours of the head indicate the socio-demographic group, while the colours of the bodies indicate households. Clusters during home-time interactions are dominated by body colours (regardless of head colours). Clusters during work-time interactions are dominated by the head colours, regardless of the body colours, signifying the coming together of individuals from different locations during working hours. Some home-time or work-time interactions can occur between people not in the same group, which is why the people in the clusters are not strictly all the same colour.

The goal of this study is to develop a model that considers the following: i) among individuals belonging to the same characteristic group (such as age groups), contacts are neither completely random nor entirely non-random but rather semi-random, ii) individuals within the same group might have distinct sets of potential and actual contacts at home compared to those at work or school, and iii) the contacts between members of different groups may vary between households and work/school, thus distinguishing the interactions within/between groups at home from those in work/school settings.

## 2 Methods

### 2.1 Group-structured contacts with semi-random mixing

For a group-structured contact with semi-random mixing, the number of contacts, average cluster sizes, average external connections per person at home and at work, and daily probability of moving within and between group interactions are applied to individuals in the same group. Some studies have enabled the estimation of average contacts within and between age groups, for example, the social contact data such as CoMix [12] and the POLYMOD [13]. The framework in this study can benefit from such data to incorporate the different contributions of different groups, to develop a group-structured epidemic model suitable for modelling risk-structured disease transmission to capture recurrent group-switch behaviour and semi-random mixing, and to distinguish between population interconnectedness during home-time interactions and those at work/school.

### 2.2 Recurrent group-switch epidemic model

We assume that each individual is assigned to a specific household, where intra/inter-group connections may be different from those in the workplace and schools. At the beginning of each time-step (one day), the family group disintegrates temporarily with each member of the group *g* moving out with a certain probability, *α*^*g*^, to interact at their usual places of activities. At the end of the workday, work/school groups disintegrate with each individual returning to the household group. This process repeats each day, giving rise to the recurrent switch between the household group and the work-time group as described by the schematic diagram in Figure 1.

A model of transmission is formulated for a group *g*, which is then generalised for any group in the population. Hence, at each time step, an individual in any group *g* decides to move out of the household with probability *α*^*g*^ or remain within the household with probability (1 − *α*^*g*^). Individuals who do not move out of the household at any time step can be infected within the household with the probability of transmission within the households; however, individuals who move out can be infected during work-time interaction with the probability of transmission during work-time interaction. This will include different phases through which a person can be infected, similar to the one in the literature [11]. The novelty in the transmission term here is that it focuses on a particular group, and may be different for different groups. Considering the possibility of being infected at home and outside the home, the probability that a susceptible person of the group *g* is infected at time *t* by the member(s) of the group 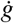 can be extrapolated from previous study [11].

The total force of infection on any group *g* from all groups in the population (see the Supplementary Information for detailed formulation) is therefore written as:

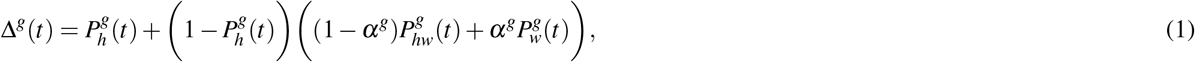

where the force of infection for group *g* within the household is given by:

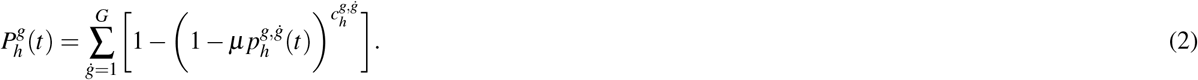

On the other hand, the force of infection at work/school is given as:

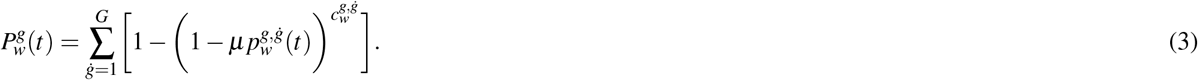

Here, 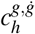 and 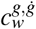 are the number of contacts made by a member of group *g* with people in group 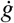 within the household and work/school per unit of time respectively, and *µ* is the per contact probability that transmission will occur given that the contact is with an infected person.

The force of infection from any group group 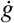 to any group *g* within households before movement to work/school, within households after movement to work/school, and at work/school are respectively given as:

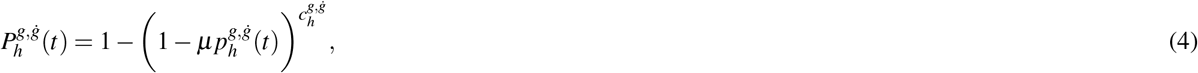

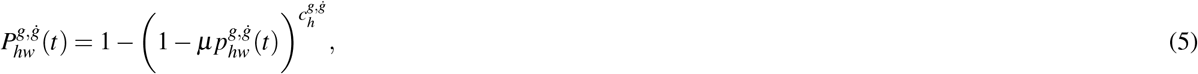

and

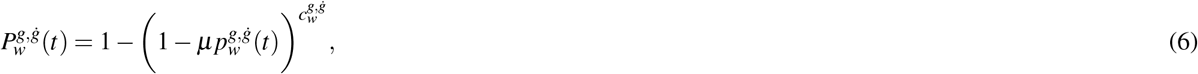

where the per-contact probability that a member of group *g* would meet with an infected member of any group 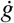 within households before movement to work/school, within households after movement to work/school, and at work/school are respectively given as:

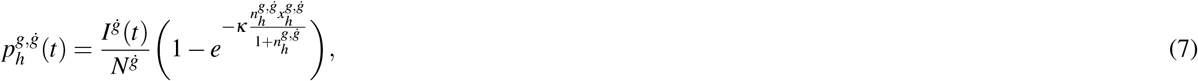

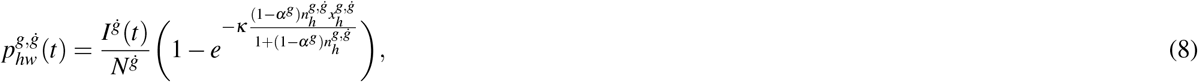

and

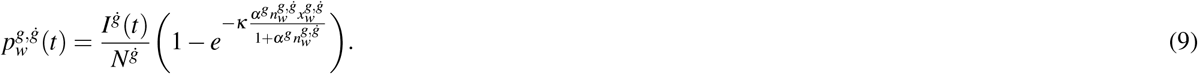

In this model, 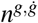 is the average number of cluster neighbours of an individual in group *g* who are members of group 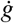, and 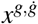 represent the average external connections per individual in group *g* who are members of group 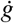, and *κ* is a population coupling parameter [10]. These terms are explicitly written because their actual values may vary depending on the group being modelled and the availability of data. The size of work clusters may vary by group; for example, office cluster sizes may differ from school cluster sizes. Similarly, average external connections may vary across groups. Here, the subscripts *h, hw*, and *w* are used to distinguish between interactions within households before movement to work/school, within households after movement to work/school, and at work/school respectively.

The force of infection for any group *g* (equation (1)) represents all infections on group *g* contributed from all groups in the populations, both at home and at work/school during home-time and work/school-time interactions. It can be used to write the equations representing the dynamics of disease transmission for the “Susceptible-Exposed-Infected-Recovered” (SEIR)-type infection represented in Figure 2, where members of group *g* who are susceptible (*S*^*g*^) become infected with a probability Δ^*g*^(*t*) and remain exposed (*E*^*g*^) until they become infectious (*I*^*g*^). During the latency period, the exposed people have a daily probability *σ* of becoming infectious. Infectious people can transmit infection to susceptibles until they are removed/recovered (*R*^*g*^) with a daily probability *γ* during which they cannot contribute to infection and cannot be re-infected.

**Figure 2.**
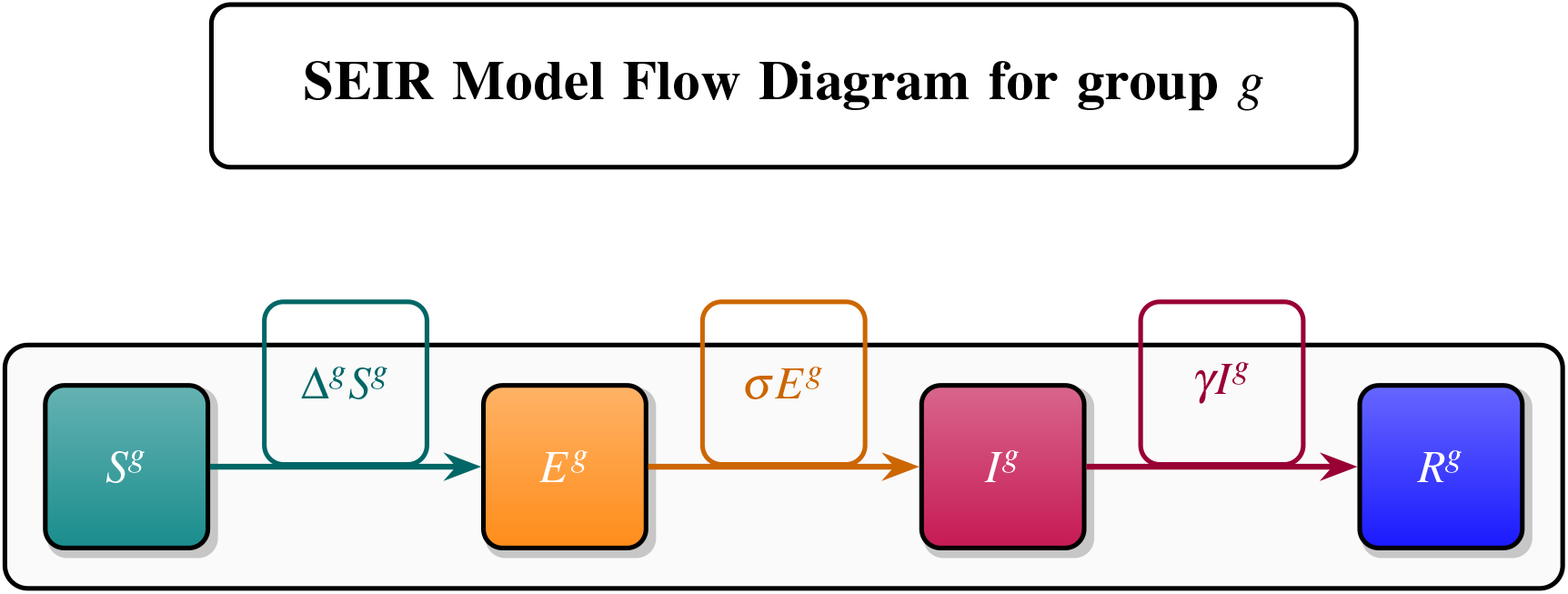
Schematic of the SEIR model for group *g*, showing Susceptible (*S*^*g*^), Exposed (*E*^*g*^), Infectious (*I*^*g*^), and Recovered (*R*^*g*^) compartments. Transitions occur at rates Δ^*g*^*S*^*g*^ (infection), *σ E*^*g*^ (progression), and *γI*^*g*^ (recovery).

The discrete-time equations for the SEIR-type infection for any group *g* are then written as:

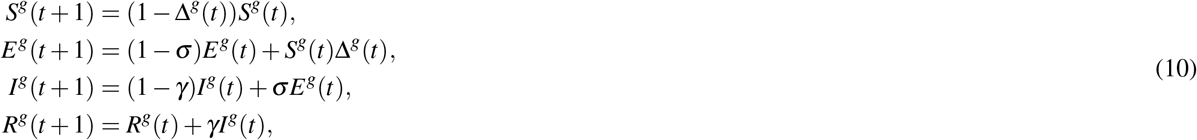

where *S*^*g*^(*t*), *E*^*g*^(*t*), *I*^*g*^(*t*), *R*^*g*^(*t*), are the numbers of the susceptible, exposed, infectious, and recovered (removed) individuals at time *t* respectively, and *S*^*g*^(*t*) + *E*^*g*^(*t*) + *I*^*g*^(*t*) + *R*^*g*^(*t*) = *N*^*g*^, where *N*^*g*^ is the number of individuals in group *g*.

### 2.3 Derivation of the Reproduction Number

We computed the time-dependent reproduction number, ℛ_t_, and basic reproduction number, ℛ_0_, using the NGM formula for the discrete-time model [14, 15]. For the *SEIR*-type model (10), the infected compartments (E, I) are used to derive the reproduction number (see the Supplementary Information).

### 2.4 Source-to-Sink reproduction number

One of the most crucial motivations for developing mathematical models in epidemiology is to understand the mechanisms needed to control or prevent outbreaks that threaten lives and economic development. Although ℛ_0_ can be useful when designing control strategies during outbreaks, the overall ℛ_0_ does not give insights into the individual contributions of each group in the spread of pathogens [16].

In heterogeneous populations, there are other methods for quantifying and defining the threshold quantity for epidemics to spread. One such method is the Type-reproduction number (*T*) [16, 17] which is defined as the expected number of secondary cases in individuals of a certain type (e.g., group 1) resulting from an infected member of their type in a totally susceptible population. These cases can result from direct transmission between members of the same type or through chains of transmission generated from an individual of the said type that passes through individuals of other types.

The model structure in this study contains parameters for interaction within/between groups, both at home and at work, so it is possible to define three quantities similar to the ℛ_0_ to gain insight into the contribution of each group in the epidemic process. These quantities are the *source*, the *sink*, and the *source-to-sink* reproduction numbers. They are used to keep track of the infection’s origin group (herein referred to as the source) and the affected group (the sink). The *Source-to-sink* reproduction number, denoted here by 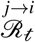 is derived from the *F*_*i j*_ element of the NGM which quantifies the average number of new infections in group *i* (the sink) resulting from infected member of group *j* (the source) during their infectious period. This measures the risk of transmission within and between groups and allows a better understanding of how appropriate, focused interventions can be implemented. Individuals from certain groups may be more susceptible to infection from other groups than others, and more generally, some groups may have a greater influence on the overall dynamics of infection in the population than others. Because heterogeneous populations are characterised by heterogeneous mobility and other characteristic structures, this approach is a tool to assess the risk of disease transmission both within and between groups in the population. The *source* reproduction number, denoted here by 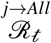, is the average number of new infections that can be created in the population from an infected individual of the source group *j*. Finally, the *sink* reproduction number, denoted here by 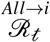, is the average number of new infections that can be created from all groups in the sink group *i*.

We define the following:

- The time-dependent *source to sink* reproduction number of group 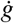 (source) on group *g* (sink) is the *F*_*i*, *j*_ element of the matrix (44) in the Supplementary Information (where *g* corresponds to *i* and 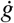 corresponds to *j*) computed as:

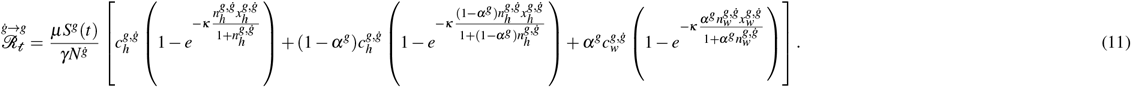
- The time-dependent *source* reproduction number of group 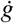 (source) on all other groups is the sum of the 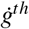 column of the matrix (44) in the Supplementary Information computed as:

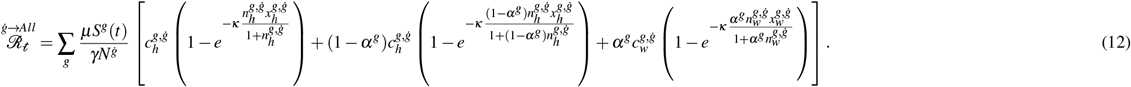
- The time-dependent *sink* reproduction number of group *g* (sink) is the sum of the *g*^*th*^ row of the matrix (44) in the Supplementary Information computed as:

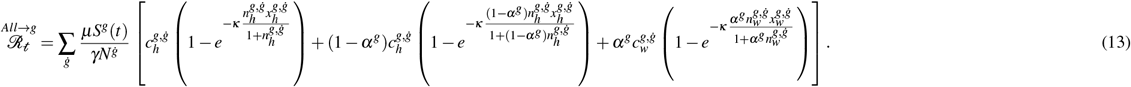

The reproduction numbers derived using this framework rely primarily on two fundamental epidemic parameters: the transmission probability, *µ*, and the recovery probability, *γ*. However, this framework reveals how social and demographic parameters impact its value. These reproduction numbers could serve as important indicators for public health officials and policymakers in formulating effective targeted strategies for pathogen control and containment.

From the analytical expressions of these quantities, the *source* and *sink* reproduction numbers give information that the basic reproduction number may not explicitly give, as they reveal the contribution to and from each group in the pathogen propagation. Also, the *source-to-sink* reproduction number gives additional information that is not readily available using the basic, the *source*, and the *sink* reproduction numbers, as it gives the information about the impact of each group on the other groups in the spread of pathogens.

### 2.5 Numerical simulation

We wanted to simulate a COVID-19-like infection; therefore, we sourced parameter values that closely match those of COVID-19. Publicly available UK social contact (POLYMOD) data [18] that shows the average number of contacts per day, within and between age groups, is used. To match the age groups, real-time infection hospitalisation and death risk of COVID-19 pandemic in England [19], the mixing data have been extracted for age groups (6–24, 25–44, 45–54, 55–64, 65–74, and 75 plus). The data have been extracted according to these specifications: (i) physical contact, (ii) contact duration of more than 15 minutes to reflect the contact duration required for the transmission of COVID-19 [20], (iii) all contact types (including holidays and weekends), and (iv) all genders. Two sets of data with these specifications were extracted: one for household mixing, representing *c*_*h*_ across age groups, and the other for non-household (work/school) mixing, representing *c*_*w*_ across age groups. We assumed that everyone will make contact with everyone else within each household, so that the household size setup *n*_*h*_ is the same as the household social contact network. Based on the class sizes of 26.4 and 22.5 in primary and secondary school in the UK, respectively [21], the average cluster members at work across groups is assumed to be ten times the work contacts. We assumed that external connections across groups at home, *x*_*h*_, is twice the number of contacts across groups at home, while the average external connections at work, *x*_*w*_, is assumed to be the same as the number of contacts across groups. The summary of the contact network data is in Figure 2 (Supplementary Information), and the rest of the parameters in Table 1.

**Table 1.**
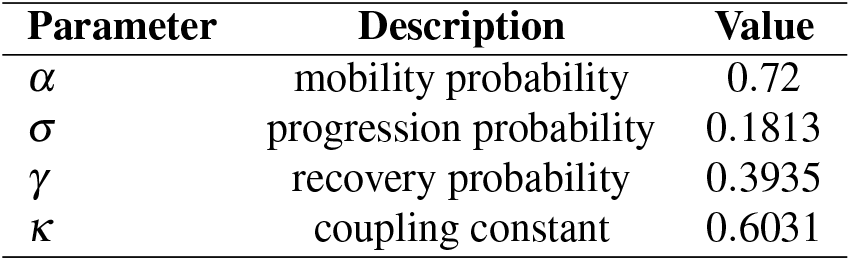
Model parameters. (Notes: *σ* and *γ* shown are probabilities per time step obtained from rates via *P*_*r*_ = 1 −*e*^−*rt*^.)

Due to the difficulty in obtaining the actual data on these parameters, the data used here is solely for the purpose of demonstrating the framework alone, while stimulating research involving the need to obtain the actual data. Other parameters are: Probability of leaving households to work/school is uniformly set for all age groups as *α* = *Average Work Days in Simulation Days/Maximum Simulation Days*. Simulating for 150 days gives *α* = 0.72, assuming that people go out to work/school for five days each week. The values of the basic reproduction number ℛ_0_ = 2.7, the rate of progression from being exposed to infectious *σ* = 0.2, and the rate of recovery *γ* = 0.5 were sourced from the literature [22]. Since *σ* and *γ* are rates, we converted them using a similar approach in the literature [4] (page 8) as *Pr* = 1 − *e*^−*rt*^, where *r* is the rate to be converted and *t* is the time step. This conversion gives the recovery probability *γ* = 0.3935, and the progression probability, *σ* = 0.1813. The coupling parameter as fitted in the literature [10] is *κ* = 0.6031. Using these parameter values, we computed the per-contact probability of infection *µ* = 0.5141 so that the relationship in equation (46) in the Supplementary Information gives ℛ_0_ = 2.7. All age groups are uniformly populated with *N* = 5000 individuals each, exposed, *E*, and recovered, *R* individuals are uniformly set to zero across all age groups, and similarly, except for the age group 6–24, where one person is initially set as infected, the number of infectious individuals is set to zeros in all age groups. Except where indicated, these parameter values are used for simulation and referred to as the baseline values. We computed the number of COVID-19 hospitalisations using data from the literature [19] as extracted and tabulated in Table 2. These numbers are computed from the number of infections generated in the numerical simulation of the *SEIR*-type model. For example, using the number of exposed individuals who progress to the infectious state at any time, we estimated new hospitalisations that would be expected from this number as *σ E*^*g*^ *×* IHR.

**Table 2.**
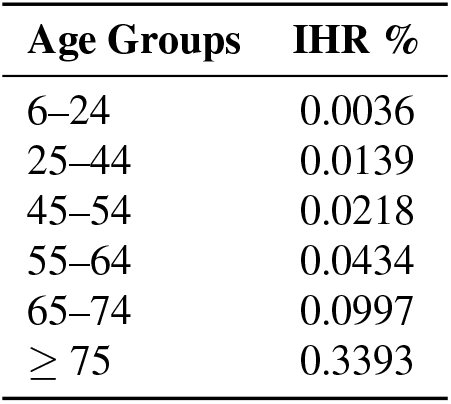
Infection Hospitalisation Risk (IHR) by Age - COVID-19.

| Age Groups | IHR % |
| --- | --- |
| 6–24 | 0.0036 |
| 25–44 | 0.0139 |
| 45–54 | 0.0218 |
| 55–64 | 0.0434 |
| 65–74 | 0.0997 |
| $\geq 75$ | 0.3393 |

Equations (46) in the Supplementary Information and (11)-(13) are used to numerically compute and compare the different reproduction numbers. The aim is to visualise the impacts and vulnerability of each age group in generating new infections in the population.

#### 2.5.1 Effects of change in baseline parameter values

We implemented non-pharmaceutical interventions by choosing a set of parameters and making a change in their baseline values, one at a time, while keeping the rest of the parameters as they are, to run simulations. The intervention strategies implemented are i) reduce work/school cluster size (*n*_*w*_) and external connections (*x*_*w*_) for ages 6–24, ii) reduce work/school cluster size (*n*_*w*_) and external connections (*x*_*w*_) for all ages, iii) reduce household and work/school external connections (*x*_*h*_, *x*_*w*_) for all ages, and iv) reduce mobility (*α*), work/school cluster size (*n*_*w*_) and external connections (*x*_*w*_) for all age groups. All interventions are implemented by reducing the baseline values of the targeted parameters in each strategy by 50%.

To analyse the effect of work/school closure for different social connectivities, we changed the baseline household and work/school external connections (*x*_*h*_, *x*_*w*_) along with the mobility probability (*α*). The focus is to see whether or not increasing mobility to work/school would always increase the epidemic final sizes, particularly considering that different populations may have different connectivity structures. We used **Algorithm 1** (Supplementary Information) to compute the final sizes for each combination of modified parameters *α* and *x*_*h*_*/x*_*w*_, using different percentages (0–100%) of the baseline values.

## 3 Results

Comparative plots showing the number of infected individuals for each age group are in Figure 3. Although the population sizes, mobility rates, and all epidemiological parameters are set equal across all age groups, differential peak timing and magnitude reflect variations in contact rates across age groups. Moreover, the initial infection introduced in age group 6–24 eventually spread to other groups, demonstrating how this framework simulates infection spreads through age-structured mixing patterns. The highest peak result is in the age group 6–24, followed by older people aged 75 and above. This confirms the mixing network in Figure 2 (Supplementary Information) where school-age 6–24 mixes most within themselves outside the households, reflecting high contacts at schools. Older people have the highest contact at home, with their highest contacts being the school age, most likely through strong interaction between grandparents and their grandchildren.

**Figure 3.**
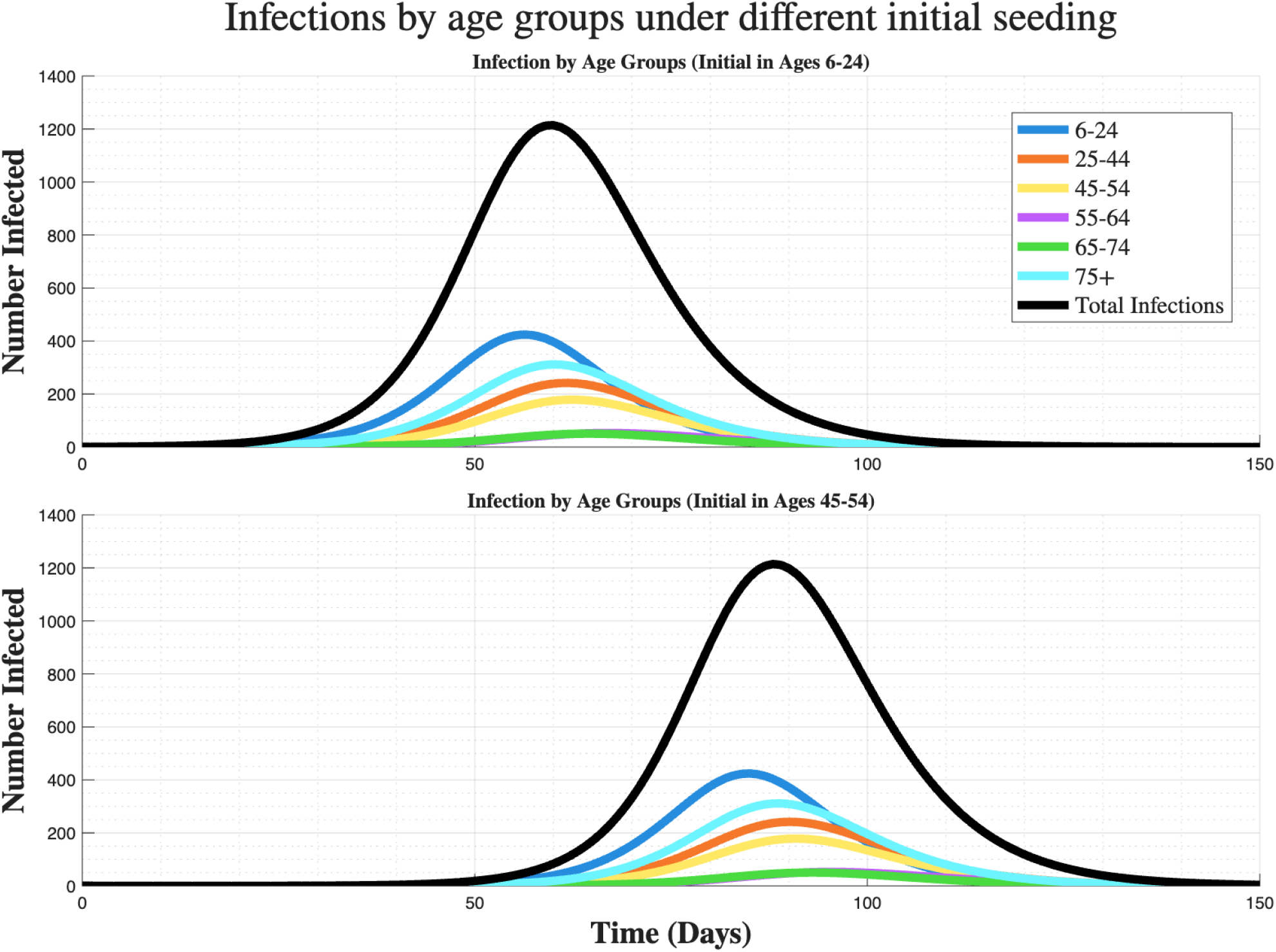
Comparison of the total number of infections over time across different age groups (6–24, 25–44, 45–54, 55–64, 65–74, 75+ years), showing the epidemic spread with the total infections aggregated across all age groups. Top: one initial infection in ages 6–24; Bottom: one initial infection in ages 45–54. Coloured lines show infections by age group; the black line is the total across groups.

Figure 3 (Top) shows a pronounced peak in total infections around day 60, while starting infection in age group 45–54 (Bottom) shows the peak around day 85, with age group 6–24 showing the highest initial infection, before a rapid decline in both cases. This suggests that younger age groups are drivers of early transmission, likely due to higher social connectivity (e.g., in school and at home). The older age group (75+ years) shows a lower infection growth as compared to those aged 6-24 years, but shows a higher infection peak as compared to other age groups. It highlights the difference in the speed of the outbreak depending on the group of initial infection.

Figure 4 illustrates the effect of interventions starting around day 30. All intervention scenarios show a marked decrease in peak infections compared to the baseline. This suggests that a 50% reduction in key transmission parameters can flatten the curve, reducing the peak number of infected individuals. Allowing mobility to work/school while reducing external connectivity at home and at work is more effective in reducing the peak height of infection.

**Figure 4.**
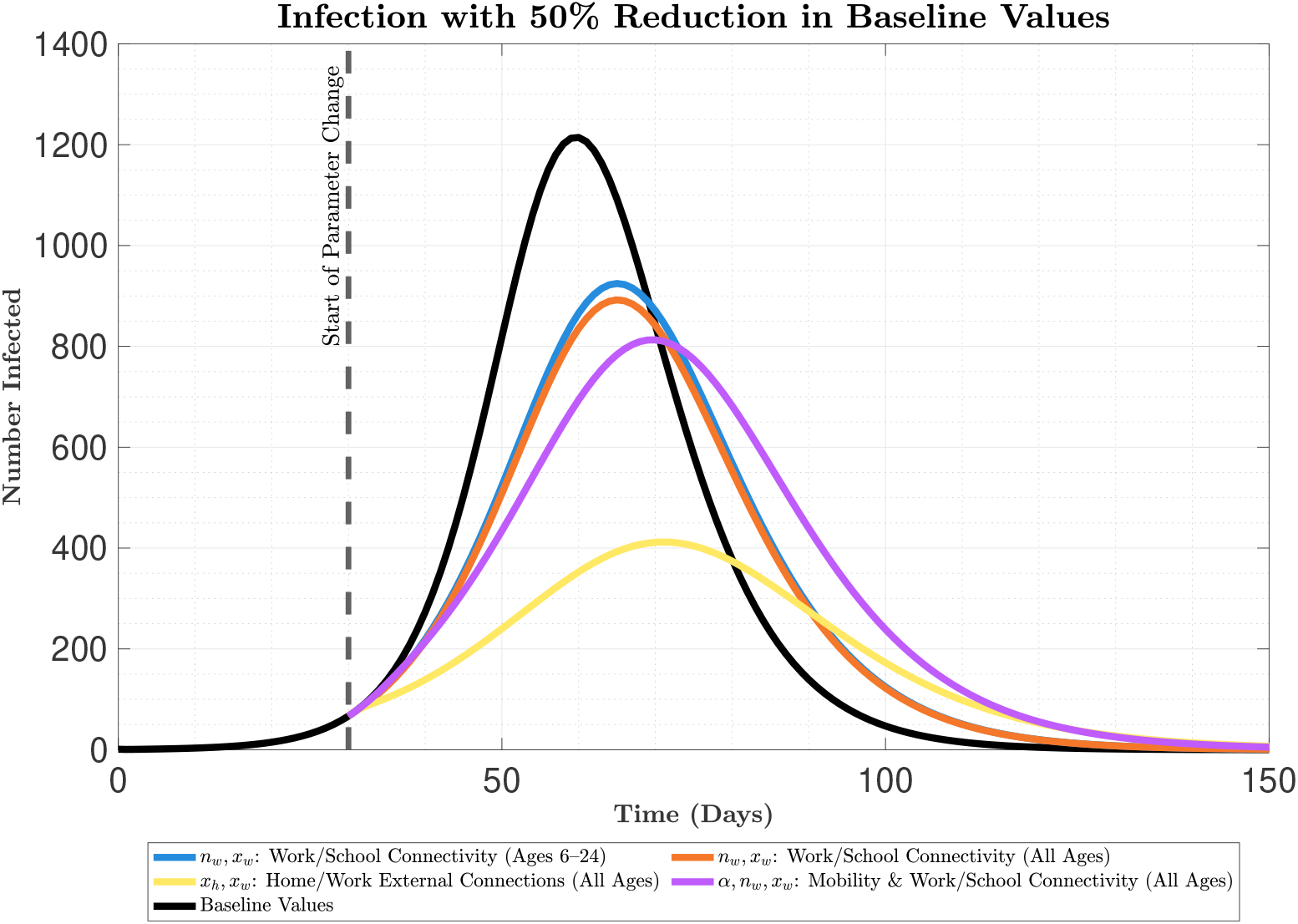
Infection dynamics with a 50% reduction in baseline values, comparing different non-pharmaceutical intervention (NPI) scenarios (work/school connectivity for ages 6–24, all ages, home/work external connections, and mobility with work/school connectivity) against baseline values, with the intervention start marked at 30 days.

The comparison of reproduction numbers in Figure 5 shows that the overall basic reproduction number is approximately 2.7, implying that each infected person in the population is likely to infect, on average, 2.7 other people before recovery. However, the *Source* and *Sink* reproduction numbers reveal that those aged above 44 years have their *Source* reproduction numbers of less than one. The age group 6-24 years has the highest *Source* and *Sink* reproduction numbers, exceeding the threshold of 1, indicating a high potential for sustained transmission from this group. The *Source* and *Sink* reproduction numbers for ages 6–24 years show that this age group will create more infection than infection would be created in them by other groups. This result is similar for age groups 25–44 years. Those in the age group 75+ years have a higher *Sink* reproduction number compared to their *Source* reproduction number (more likely to be infected than infect others). A similar trend is seen in age groups 45–54, 55–64, and 65–74 years. Thus, the source and sink reproduction numbers indicate that the 6-24 year age group acts as a significant source of infection, while older groups are more likely to be sinks, receiving infections from younger populations. This demonstrates the need for risk-structured modelling, as in this study, to inform interventions during outbreaks.

**Figure 5.**
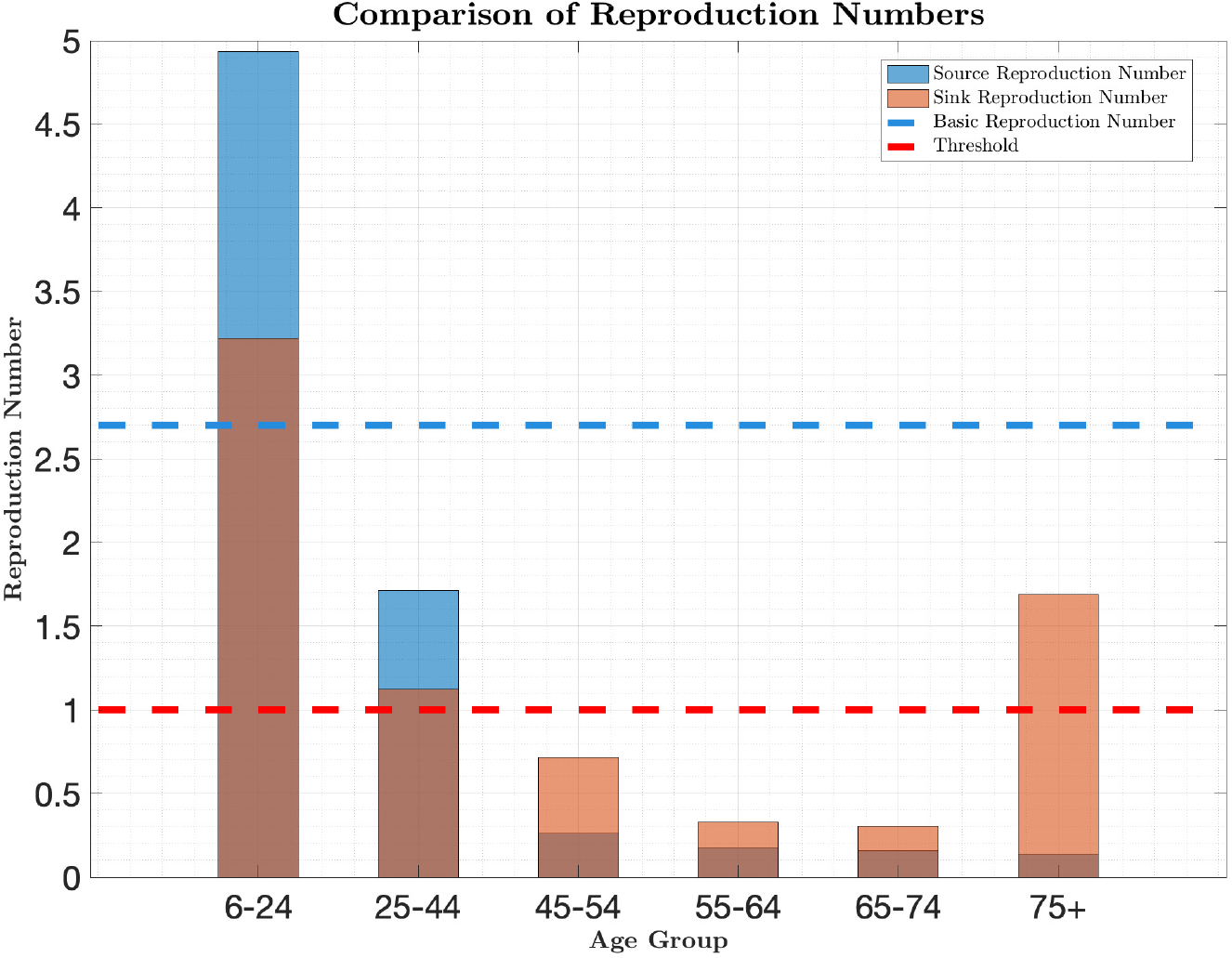
Age-specific reproduction numbers decomposed into source and sink components, with the basic reproduction number ℛ_0_ (dashed blue line) and epidemic threshold (dashed red line).

The Source-to-Sink reproduction numbers heatmap in Figure 6 indicates that the highest transmissions occur within and from the 6–24years age group, with transmission to other groups decreasing with age. This age-specific transmission pattern supports the results in Figure 5, which indicate that school-aged groups (6–24years) are a key source of transmission, and the highest transmission is within the age group than with others. Interventions targeting this group could have the greatest impact on reducing overall transmission.

**Figure 6.**
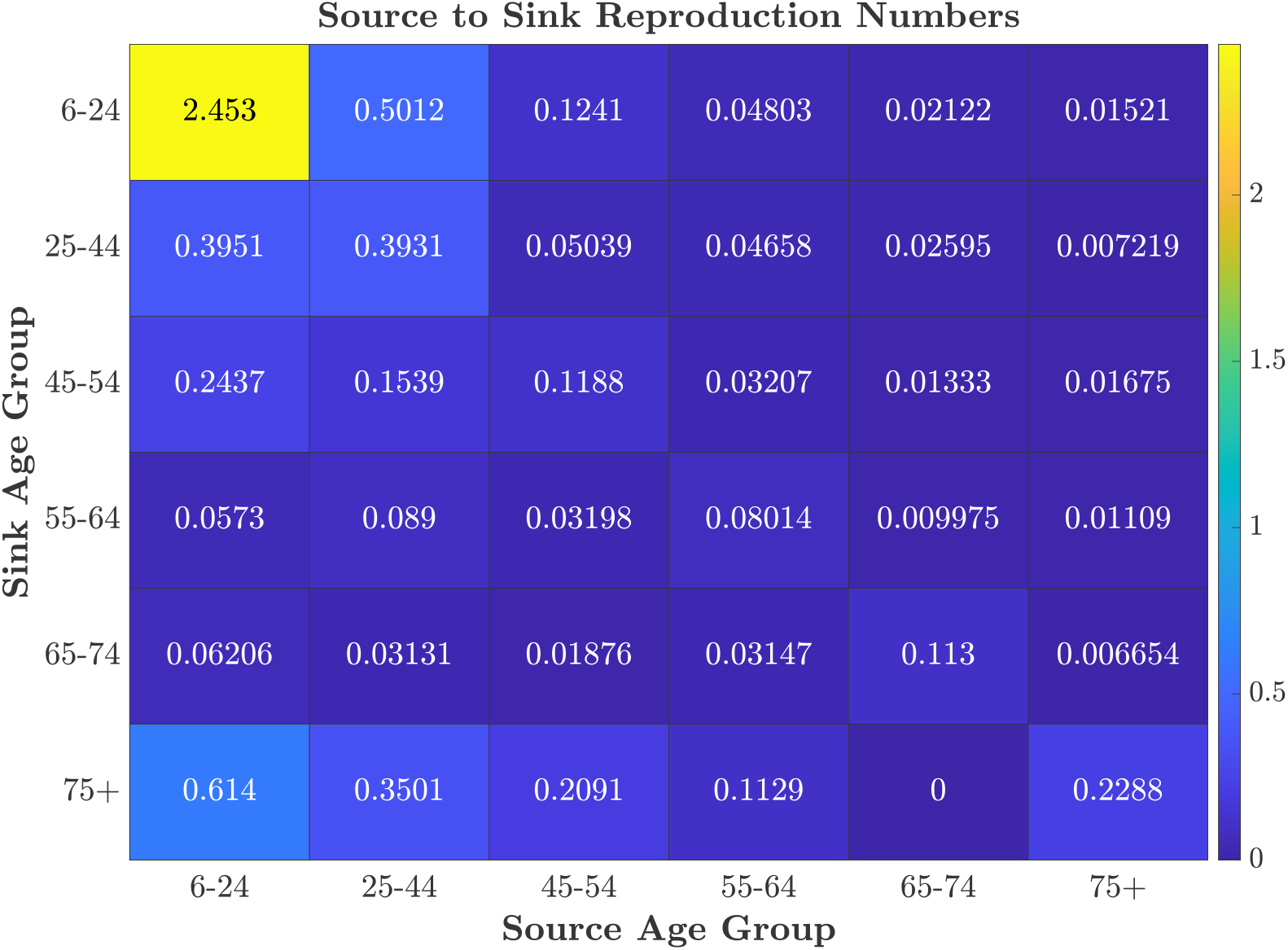
Source-to-Sink reproduction number matrix quantifying transmission from age group *g* (source) to 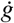 (sink). The diagonal represents within-group transmission, while off-diagonal elements show cross-group transmission. Bright regions indicate high transmission pairs.

The results in Figures 7 and 8 highlight that age groups 75+ years experience the highest hospitalisation, particularly under baseline conditions.

**Figure 7.**
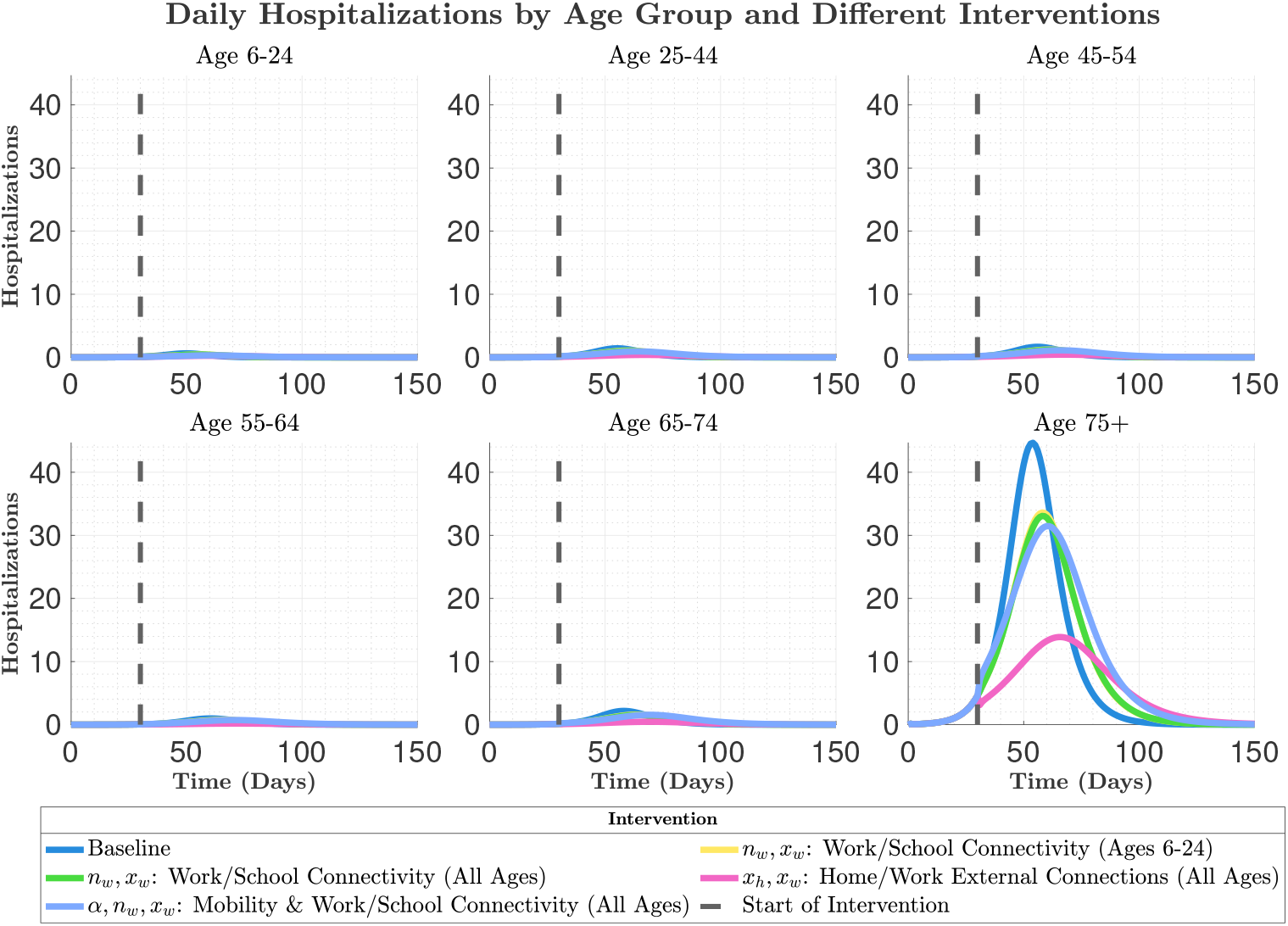
Daily hospitalisation by age group over time across different intervention scenarios. The interventions are implemented by *±*50% change in the baseline parameter values.

**Figure 8.**
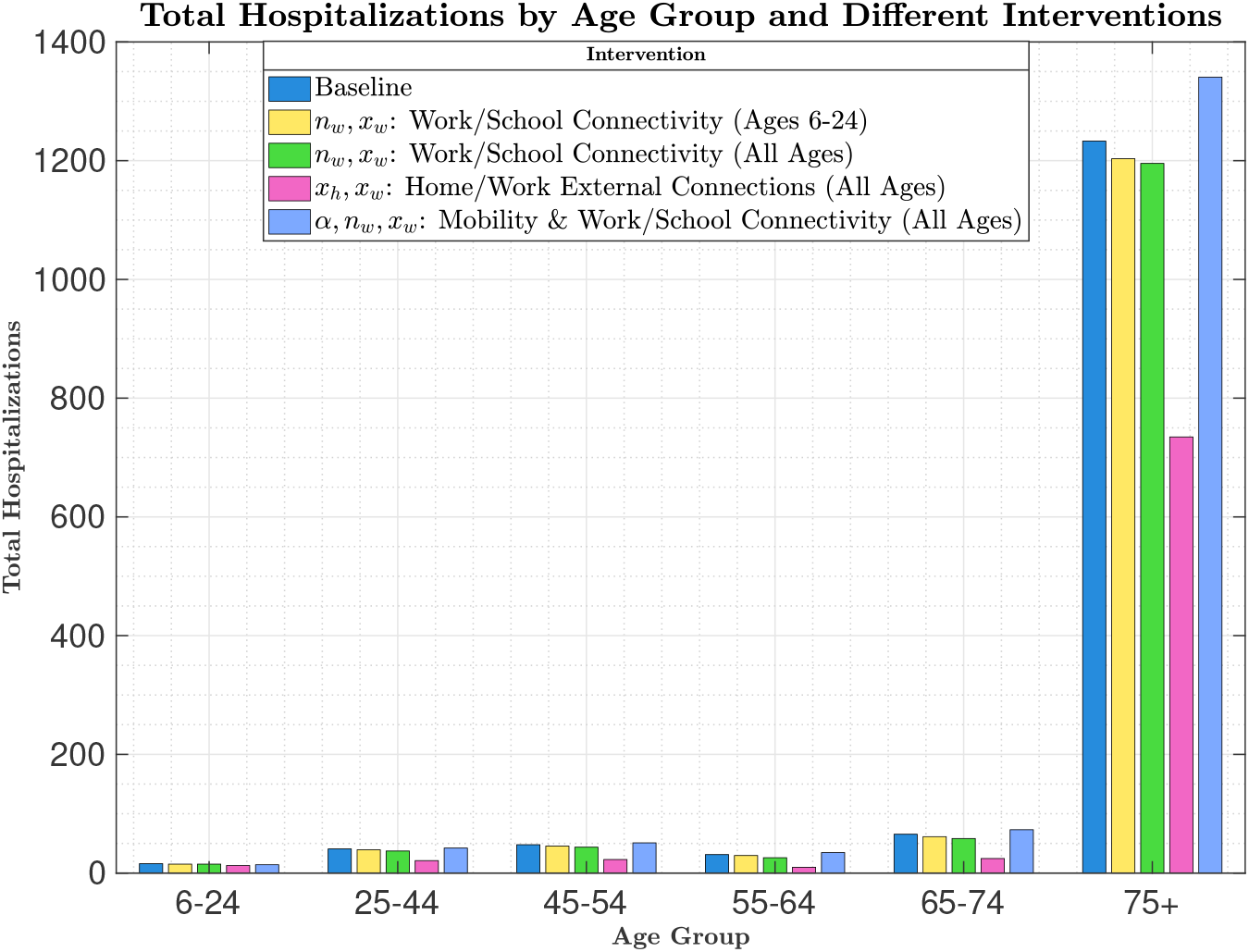
Bar plot of total hospitalisations by age group (6–24, 25–44, 45–54, 55–64, 65–74, 75+) across different NPI scenarios. The interventions are implemented by *±*50% change in the baseline parameter values.

Figures 9 and 10 demonstrate the effect of increasing mobility for each age group with varying external connections at work and within the households, respectively. Figure 9 shows no effect of work/school closure on age groups 6–24 years if the average external connections at work *x*_*w*_ for all age groups are above 40% of the baseline values. It shows that reducing mobility of all age groups to below 50% of the baseline values, while the average external connections at work *x*_*w*_ for all age groups are below 40%, would increase final sizes in all age groups.

**Figure 9.**
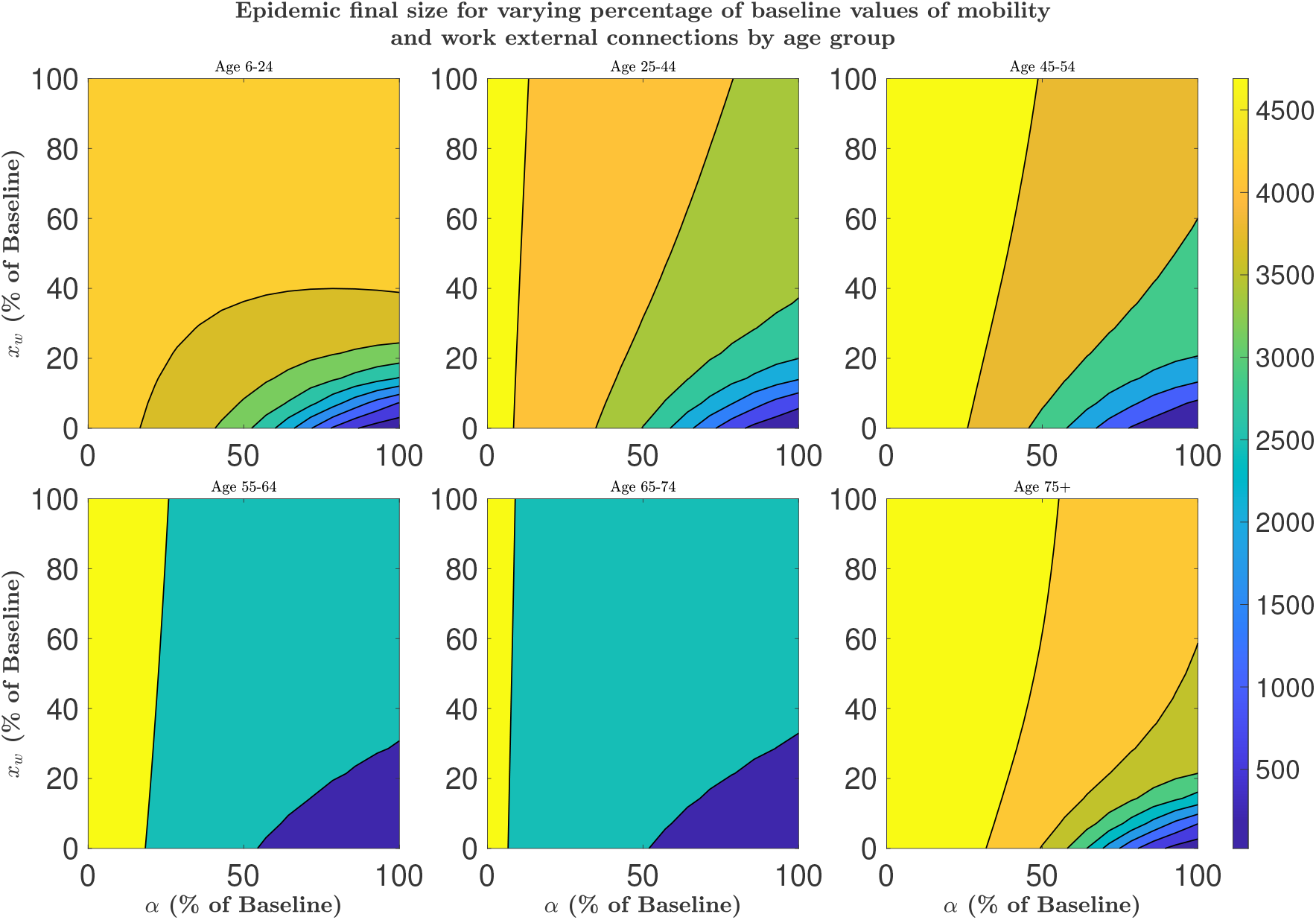
Contour plots showing the epidemic final size as a function of varying percentages of baseline mobility (*α*) and external connections at work/school (*x*_*w*_) across age groups (6–24, 25–44, 45–54, 55–64, 65–74, 75+). Parameters (*α*) and (*x*_*w*_) are scaled from 0% to 100% of their baseline values. Lighter colours correspond to higher epidemic final size.

**Figure 10.**
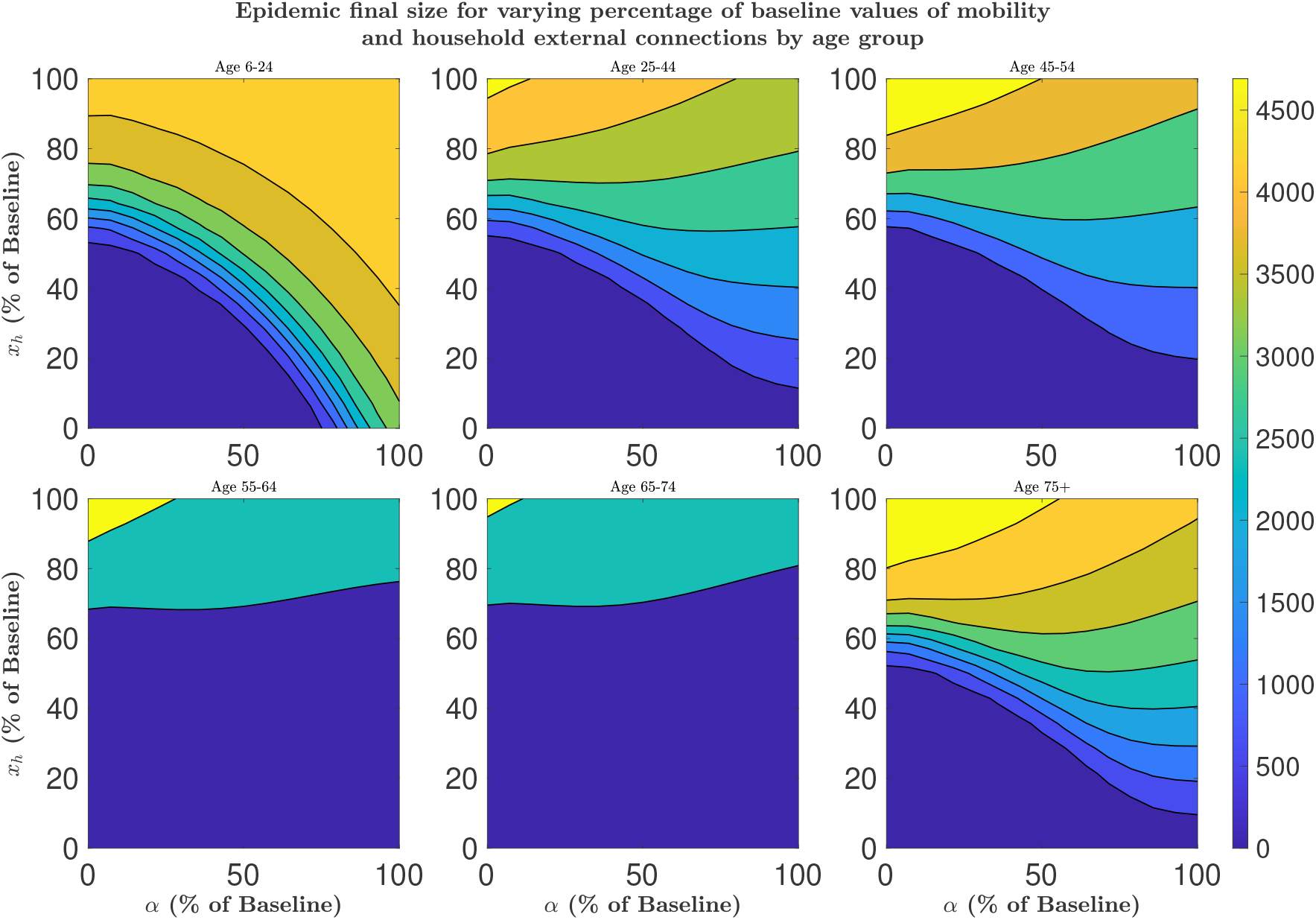
Contour plots showing the epidemic final size as a function of varying percentages of baseline mobility (*α*) and external connections at home (*x*_*h*_) across age groups (6–24, 25–44, 45–54, 55–64, 65–74, 75+). Parameters (*α*) and (*x*_*h*_) are scaled from 0% to 100% of their baseline values. Lighter colours correspond to higher epidemic final size.

Figure 10 show that, if the household external connection, *x*_*h*_ representing inter-house interactions are below 15% of the baseline values, then increase/decrease in mobility does not have visible impact on the final sizes in all age groups except for ages 6-24 which shows increase in the final size for mobility above 70% of the baseline value. Household external connections above 60% of the baseline values would mean that increasing mobility to work/school reduces the final sizes of age groups 25–44, 45–54, and 75+ years. From these results, change in mobility would impact the final size in ages 55–64 and 65–74 only when the household external connection, *x*_*h*_, is above 70% of the baseline values.

Figure 11 is a contour plots that show the total epidemic final size. The plots show how final size changes with baseline mobility (*α*) and external connection parameters *x*_*h*_ and *x*_*w*_. The focus is on the ranges of *x*_*h*_ and *x*_*w*_ within which variations in mobility (*α*) result in either an increase or a decrease in the final size.

**Figure 11.**
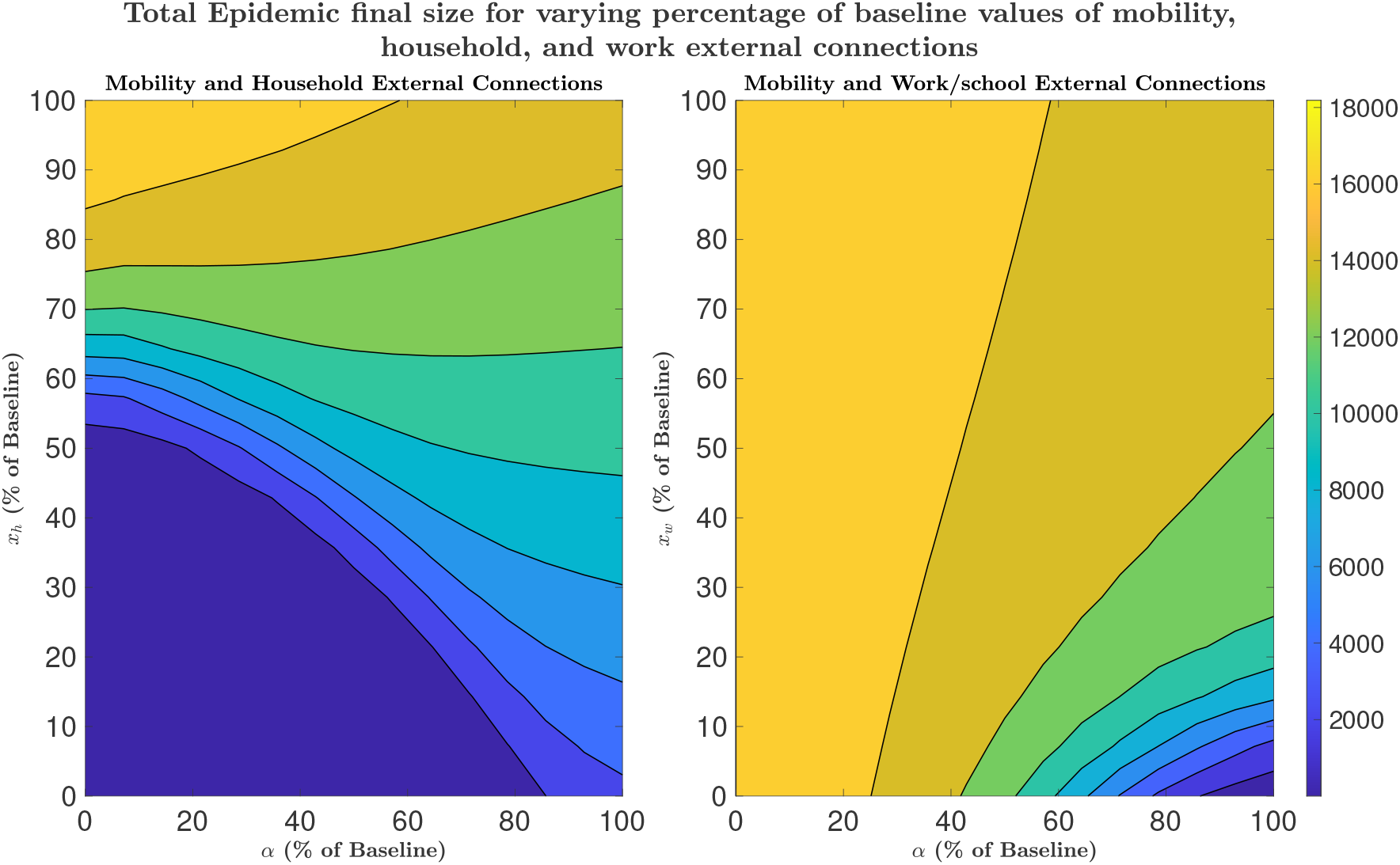
Contour plots of total epidemic final size across all age groups as a function of varying percentages of baseline mobility and external connections (household and work/school), with parameters scaled from 0% to 100% of their baseline values. **Left:** Total epidemic final size across all age groups varying with percentage of baseline mobility and household external connections, (*α* and *x*_*h*_). **Right:** Total epidemic final size across all age groups varying with percentage of baseline mobility and work/school external connections, (*α* and *x*_*w*_).

When mobility (*α*) is increased above 50% of its baseline value, the final size increases (Figure 11) (left). This rise is most noticeable when *x*_*h*_ is between 55 and 70%. When mobility is increased and *x*_*h*_ is above 85% of the baseline values, the final size reduces, which is counter-intuitive.

For the relationship between *α* and *x*_*w*_ (Figure 11) (right), the final size decreases when mobility (*α*) exceeds 25% for all values of *x*_*w*_. This effect is very strong when *x*_*w*_ is less than 50%. These results support other studies in the literature [23–26] which found that under certain network structures, increasing mobility would prevent the spread of infectious diseases.

## 4 Discussion

In this study, we developed and analysed a recurrent group-switch epidemic model that integrates semi-random mixing, group-structured contacts, and daily transitions between household and work/school places. The findings provide new insights into how heterogeneous contact structures and mobility patterns shape pathogen transmission, and how age-specific social/behavioural attributes in epidemic modelling can inform targeted public health interventions.

A key result is the identification of the 6–24 years age group as a primary source of transmission. Despite equal population sizes and epidemiological parameters across groups, the higher connectivity of younger individuals—typically in schools and through household interactions—produced higher and earlier infection peaks. This is in agreement with empirical evidence from the COVID-19 pandemic, where children and adolescents show higher infection, but lower disease severity [6]. In contrast, ages 75 years and above were primarily recipients of infections (“sinks”) rather than drivers, but had higher hospitalisation risks due to the severity of illness [19]. It also shows that the group from which the first infection is introduced could determine the speed of spread in the population.

The comparative analysis of NPIs highlights the importance of targeted intervention strategies. Reducing school/work between-clusters connectivity in the 6–24 years age group produced effects comparable to when this reduction is implemented across all age groups, indicating that interventions focused on school-aged populations can achieve a reduction in epidemic spread. This approach would lead to desirable results with fewer societal disruptions. This result supports earlier modelling studies suggesting that school closures, cohorting, or hybrid learning can reduce transmission during early epidemic phases [22]. However, the model also highlights the limitations of work/school-based mobility restrictions as a stand-alone intervention strategy: reducing mobility without curbing inter-household or inter-cluster connections increased the potential for multi-phased household-based transmission. Interventions that allow essential mobility while limiting inter-household connections (e.g., restrictions on large gatherings or inter-household mixing) are seen to be more effective in flattening epidemic curves than strictly mobility restrictions.

From the analysis of the final sizes, under some population structure, restricting mobility to work/school would increase the epidemic final size. This counterintuitive result supports previous studies [24–26], which found that in certain regimes, increasing mobility could render the population less prone to outbreaks. These findings caution against assuming strictly positive effects of restricting work/school-based mobility and highlight the importance of tailoring NPIs to the structural properties of populations.

From a public health perspective, hospitalisation analyses reaffirm the vulnerability of older adults. Even when acting predominantly as sinks of transmission, individuals aged 75+ generated the highest number of hospitalisations, reflecting the variability in COVID-19 hospitalisation risk [19]. These results emphasise the need for integrated intervention strategies: reducing transmission in younger, high-contact groups indirectly protects older populations.

Methodologically, this work extends traditional risk-structured models by incorporating recurrent group-switch and semi-random mixing behaviours, thus capturing daily cycles of household and work/school interactions, essentially presenting a framework to allows for analysis of both household and non-household transmission more realistically. The explicit derivation of source, sink, and Source-to-Sink reproduction numbers contributes to the analytical insights for epidemic modelling in heterogeneous populations. Unlike the basic reproduction number (ℛ_0_), these measures reveal directional contributions to transmission between groups, enabling more precise evaluation of group-specific risks and intervention impacts.

While the recurrent group-switch model adds to epidemic modelling, several limitations must be acknowledged. First, the simulations relied on assumed parameterisations of cluster sizes and external connections due to the limited availability of empirical data. Although social contact surveys such as POLYMOD and CoMix [12, 13] informed baseline estimates, more detailed data on household structures, workplace sizes, and inter-household/cluster mixing would improve realism. Second, the model assumes homogeneity within age groups, neglecting behavioural, socioeconomic, and occupational heterogeneity that can substantially affect transmission dynamics even within the same population group. Similarly, adherence to NPIs, testing, and isolation strategies—critical components of epidemic control—were excluded. Analytically, the derivation of reproduction numbers becomes increasingly complex with greater stratification. While the next-generation matrix framework accommodates multiple groups, interpretation and computation may be challenging for highly granular stratifications, such as joint age-occupation-location structures. Future work could integrate data-driven approaches to calibrate parameters or explore hybrid agent-based and equation-based models to bridge tractability with realism. Although the mobility structure in this model allows the distinction between work/school-based mobility from the possible movement and interactions between households, the specific destinations-typically work/school locations that reflect the realistic origin and destination for different people, have not been included. Implementing this framework in the light of spatial metapopulation, where different locations interact during work/school, would increase the realism of this framework and help analyse the impacts of mobility in population-wide dissemination of pathogens that would lead to epidemics or global pandemics. Finally, the present framework was motivated by COVID-19-like dynamics, and children between the ages of 0–5 have been omitted in the age stratification. Its application to other pathogens with different age-specific risk structures, to include all age groups (e.g., influenza, Ebola), requires careful adaptation of contact, severity, and transmission parameters. Nevertheless, the generality of the recurrent group-switch concept makes it broadly applicable to pathogens transmitted via close contact. The recurrent group-switch framework not only improves understanding of epidemic dynamics but also provides a foundation for evaluating multi-layered, context-specific public health strategies in heterogeneous populations.

## Acknowledgements

This research is fully funded by the Institute for Global Pandemic Planning, Warwick Medical School, University of Warwick UK.

## Author contributions statement

Conceptualization: M. L. S.

Methodology: M. L. S. Supervision: A.S. & K. S. R.

Software: K. S. R. & M. L. S. Visualisation: K S. R & M. L. S.

Writing – original draft: M. L. S.

Writing – review & editing: A. S., K. S. R. & M. L. S.

## Data availability

The MATLAB code used to generate the figures is available on Zenodo https://doi.org/10.5281/zenodo.17605791

## Ethics declarations

This study focuses on the theoretical development of a mathematical model for the spread of a hypothetical infectious disease. No data was collected and analysed.

