## Supplementary Information for "Recurrent Group-switch Interactions in Heterogeneous Population Epidemic Modelling"

### 1 Methods

To formulate the recurrent group switch model, we adopt the force of infection developed in [1], which is given as:

$$FOI(t) = \beta \frac{I(t)}{N}\left(1 - e^{-\kappa \frac{nx}{n+1}}\right). \tag{1}$$

Where $\beta = \mu c$ is the transmission rate, where $\mu$ is the per-contact probability that infection will be transmitted given a contact with an infected person, and $c$ is the average number of contacts per person in the population. The term $\frac{I(t)}{N}$ is the disease prevalence in the population, while the last term, $1 - e^{-\kappa \frac{nx}{n+1}}$ is the contact saturation function that accounts for how likely it is that everyone will connect with everyone else, where $\kappa$ is a coupling constant, $n$ is the average cluster size, and $x$ is the average external (inter-cluster) connections per person in the population. Here, $I(t)$ is the number of infected individuals at time $t$, $N$ is the total population (healthy and infected). This specific formulation is only suitable for modelling disease transmission in a homogeneous population without consideration of the differences in population groups.

#### 1.1 Group-structured contacts with semi-random mixing

For a group-structured contact with semi-random mixing, the number of contacts, average cluster sizes, average external connections per person at home and at work, and daily probability of moving within and between group interactions are applied to individuals in the same group. Some studies have enabled the estimation of average contacts within and between age groups, for example, the social contact data such as CoMix [2] and the POLYMOD [3]. The framework in this study can benefit from such data to incorporate the different contributions of different groups, to develop a group-structured epidemic model suitable for modelling risk-structured disease transmission to capture recurrent group-switch behaviour and semi-random mixing, and to distinguish between population interconnectedness during home-time interactions and those at work.

Let $A = \{1, 2, 3, ...G\}$ be the set of groups in a given population, and let the number of individuals in groups $1, 2, 3, ...G$ be $N^1, N^2, N^3, ..., N^G$ respectively, where $G$ is the number of groups in the population.

To write the force of infection in any specific group, say group $g$, it is easier to first write the force of infection contributed by any group $\dot{g} \in A$ to group $g$:

$$FOI^{g,\dot{g}}(t) = \beta^{g,\dot{g}} \frac{I^{\dot{g}}(t)}{N^{\dot{g}}}\left(1 - e^{-\kappa \frac{n^{g,\dot{g}} x^{g,\dot{g}}}{1+n^{g,\dot{g}}}}\right). \tag{2}$$

Here, $n^{g,\dot{g}}$ is the average number of cluster neighbours of an individual in group $g$ who are members of group $\dot{g}$, and $x^{g,\dot{g}}$ represent the average external connections per individual in group $g$ who are members of group $\dot{g}$. These terms are explicitly written because their actual values may vary depending on the group being modelled and the availability of data. The size of work clusters may vary by group; for example, office cluster sizes may differ from school cluster sizes. Similarly, average external connections may vary across groups. The total force of infection on group $g$ is thus given as:

$$FOI^{g}(t) = \sum_{\dot{g}=1}^{G} \beta^{g,\dot{g}} \frac{I^{\dot{g}}(t)}{N^{\dot{g}}} \left(1 - e^{-\kappa \frac{n^{g,\dot{g}} x^{g,\dot{g}}}{1+n^{g,\dot{g}}}}\right). \tag{3}$$

Following a similar approach for writing a non-linear force of infection in the literatur [1? ], the force of infection on group $g$ is thus given as:

$$FOI^{g}(t) = \sum_{\dot{g}=1}^{G} \left[1 - \left(1 - \mu \frac{I^{\dot{g}}(t)}{N^{\dot{g}}} \left(1 - e^{-\kappa \frac{n^{g,\dot{g}} x^{g,\dot{g}}}{1+n^{g,\dot{g}}}}\right)\right)^{c^{g,\dot{g}}}\right], \tag{4}$$

where $c^{g,\dot{g}}$ is the number of contacts that an individual in group $g$ make with members of group $\dot{g}$.

As stated earlier, we considered a multi-phased transmission— during household and work interactions. Based on this, the probabilities that a person in any group $g \in A$ will meet with an infected individual of group $\dot{g}$ during home-time and work-time interactions are thus given respectively as:

$$p_h^{g,\dot{g}}(t) = \frac{I^{\dot{g}}(t)}{N^{\dot{g}}} \left(1 - e^{-\kappa \frac{n_h^{g,\dot{g}} x_h^{g,\dot{g}}}{1+n_h^{g,\dot{g}}}}\right), \tag{5}$$

$$p_w^{g,\dot{g}}(t) = \frac{I^{\dot{g}}(t)}{N^{\dot{g}}} \left(1 - e^{-\kappa \frac{n_w^{g,\dot{g}} x_w^{g,\dot{g}}}{1+n_w^{g,\dot{g}}}}\right). \tag{6}$$

Using equations (5)- (6), the force of infection for group $g$ within the household is given by:

$$P_h^{g}(t) = \sum_{\dot{g}=1}^{G} \left[1 - \left(1 - \mu p_h^{g,\dot{g}}(t)\right)^{c_h^{g,\dot{g}}}\right]. \tag{7}$$

On the other hand, the force of infection at work/school is given as:

$$P_w^{g}(t) = \sum_{\dot{g}=1}^{G} \left[1 - \left(1 - \mu p_w^{g,\dot{g}}(t)\right)^{c_w^{g,\dot{g}}}\right]. \tag{8}$$

Where $c_h^{g,\dot{g}}$ and $c_w^{g,\dot{g}}$ are the number of contacts made by a member of group $g$ with people in group $\dot{g}$ within the household and work/school per unit of time respectively, and $\mu$ is the per contact probability that transmission will occur given that the contact is with an infected person.

#### 1.2 Recurrent group-switch epidemic model

A model of transmission is formulated for a group $g$, which is then generalised for any group in the population. Hence, at each time step, an individual in any group $g$ decides to move out of the household with probability $\alpha^g$ or remain within the household with probability $(1-\alpha^g)$. Individuals who do not move out of the household at any time step can be infected within the household with the probability of transmission within the households; however, individuals who move out can be infected during work-time interaction with the probability of transmission during work-time interaction. This will include different phases through which a person can be infected, similar to the one in the literature []. The novelty in the transmission term here is that it focuses on a particular group, and may be different for different groups. Considering the possibility of being infected at home and outside the home, the probability that a susceptible person of the group $g$ is infected at time $t$ by the member(s) of the group $\dot{g}$ can be extrapolated from previous study []. This probability for transmission interactions between households and non-households is given by:

$$\Delta^{g,\dot{g}}(t) = P_h^{g,\dot{g}}(t) + \left(1 - P_h^{g,\dot{g}}(t)\right)\left((1-\alpha^g) P_{hw}^{g,\dot{g}}(t) + \alpha^g P_w^{g,\dot{g}}(t)\right), \tag{9}$$

here $P_h^{g,\dot{g}}(t)$ is the probability of being infected at home before mobility, $(1-\alpha^g)P_{hw}^{g,\dot{g}}(t)$ is the probability of being infected at home having not left the house at time $t$, and $\alpha^g P_w^{g,\dot{g}}(t)$ is the probability of being infected at time $t$ at work/school. Each of those probabilities is given by:

$$P_h^{g,\dot{g}}(t) = 1 - \left(1 - \mu p_h^{g,\dot{g}}(t)\right)^{c_h^{g,\dot{g}}}, \tag{10}$$

$$P_{hw}^{g,\dot{g}}(t) = 1 - \left(1 - \mu p_{hw}^{g,\dot{g}}(t)\right)^{c_h^{g,\dot{g}}}, \tag{11}$$

and

$$P_w^{g,\dot{g}}(t) = 1 - \left(1 - \mu p_w^{g,\dot{g}}(t)\right)^{c_w^{g,\dot{g}}}, \tag{12}$$

with

$$p_h^{g,\dot{g}}(t) = \frac{I^{\dot{g}}(t)}{N^{\dot{g}}}\left(1 - e^{-\kappa \frac{n_h^{g,\dot{g}} x_h^{g,\dot{g}}}{1+n_h^{g,\dot{g}}}}\right), \tag{13}$$

$$p_{hw}^{g,\dot{g}}(t) = \frac{I^{\dot{g}}(t)}{N^{\dot{g}}}\left(1 - e^{-\kappa \frac{(1-\alpha^g) n_h^{g,\dot{g}} x_h^{g,\dot{g}}}{1+(1-\alpha^g) n_h^{g,\dot{g}}}}\right), \tag{14}$$

and

$$p_w^{g,\dot{g}}(t) = \frac{I^{\dot{g}}(t)}{N^{\dot{g}}}\left(1 - e^{-\kappa \frac{\alpha^g n_w^{g,\dot{g}} x_w^{g,\dot{g}}}{1+\alpha^g n_w^{g,\dot{g}}}}\right). \tag{15}$$

The total force of infection for any group $g$ from all groups in the population is therefore written as:

$$\Delta^g(t) = P_h^g(t) + \left(1 - P_h^g(t)\right)\left((1-\alpha^g)P_{hw}^g(t) + \alpha^g P_w^g(t)\right), \tag{16}$$

where $P_h^g(t)$, $P_{hw}^g(t)$, and $P_w^g(t)$ are as defined in equations 7 and 8.

The force of infection for any group $g$ (equation (16)) represents all infections on group $g$ contributed from all groups in the populations, both at home and at work/school during home-time and work/school-time interactions. It can be used to write the equations representing the dynamics of disease transmission for the "Susceptible-Exposed-Infected-Recovered" (SEIR)-type infection represented in Figure 1, where members of group $g$ who are susceptible ($S^g$) become infected with a probability $\Delta^g(t)$ and remain exposed ($E^g$) until they become infectious ($I^g$). During the latency period, the exposed people have a daily probability $\sigma$ of becoming infectious. Infectious people can transmit infection to susceptibles until they are removed/recovered ($R^g$) with a daily probability $\gamma$ during which they cannot contribute to infection and cannot be re-infected.

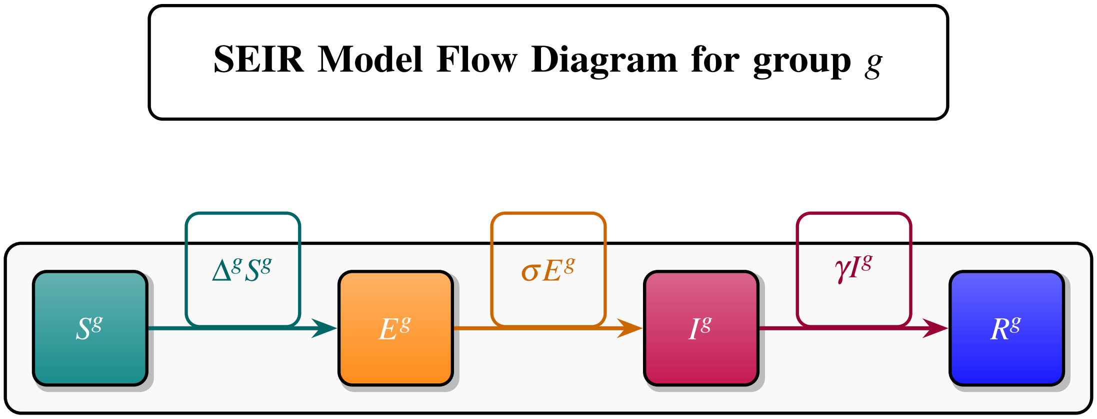


**Figure 1.** Schematic of the SEIR model for group $g$, showing Susceptible ($S^g$), Exposed ($E^g$), Infectious ($I^g$), and Recovered ($R^g$) compartments. Transitions occur at rates $\Delta^g S^g$ (infection), $\sigma E^g$ (progression), and $\gamma I^g$ (recovery).

The discrete-time equations for the SEIR-type infection for any group $g$ are then written as:

$$\begin{aligned} S^g(t+1) &= (1-\Delta^g(t))S^g(t), \\ E^g(t+1) &= (1-\sigma)E^g(t) + S^g(t)\Delta^g(t), \\ I^g(t+1) &= (1-\gamma)I^g(t) + \sigma E^g(t), \\ R^g(t+1) &= R^g(t) + \gamma I^g(t), \end{aligned} \tag{17}$$

where $S^g(t)$, $E^g(t)$, $I^g(t)$, $R^g(t)$, are the numbers of the susceptible, exposed, infectious, and recovered (removed) individuals at time $t$ respectively, and $S^g(t)+E^g(t)+I^g(t)+R^g(t) = N^g$, where $N^g$ is the number of individuals in group $g$.

#### 1.3 Derivation of the Reproduction Number

The linearised form of the equations (10–12) are:

$$\tilde{P}_h^g(t) = \mu \sum_{\dot{g}=1}^{G} \frac{c_h^{g,\dot{g}} I^{\dot{g}}(t)}{N^{\dot{g}}} \left(1 - e^{-\kappa \frac{n_h^{g,\dot{g}} x_h^{g,\dot{g}}}{1+n_h^{g,\dot{g}}}}\right). \tag{18}$$

$$\tilde{P}_{hw}^g(t) = \mu \sum_{\dot{g}=1}^{G} \frac{c_h^{g,\dot{g}} I^{\dot{g}}(t)}{N^{\dot{g}}} \left(1 - e^{-\kappa \frac{(1-\alpha^g) n_h^{g,\dot{g}} x_h^{g,\dot{g}}}{1+(1-\alpha^g) n_h^{g,\dot{g}}}}\right). \tag{19}$$

$$\tilde{P}_w^g(t) = \mu \sum_{\dot{g}=1}^{G} \frac{c_w^{g,\dot{g}} I^{\dot{g}}(t)}{N^{\dot{g}}} \left(1 - e^{-\kappa \frac{\alpha^g n_w^{g,\dot{g}} x_w^{g,\dot{g}}}{1+\alpha^g n_w^{g,\dot{g}}}}\right). \tag{20}$$

At the beginning of outbreaks, we have $t \to 0$, and $\tilde{P}_h^g(t) \to 0$, since we start with a very small fraction of the population initially infected. This approximates $1 - \tilde{P}_h^g(t) \approx 1$, to write equation (16) as:

$$\tilde{\Delta}^g(t) = \tilde{P}_h^g(t) + (1-\alpha^g)\tilde{P}_{hw}^g(t) + \alpha^g \tilde{P}_w^g(t). \tag{21}$$

Substituting for $\tilde{P}_h^g(t)$, $\tilde{P}_{hw}^g(t)$, and $\tilde{P}_w^g(t)$ in equation (21) gives:

$$
\begin{aligned}
\widetilde{\Delta}^g(t) = \mu \sum_{\dot{g}=1}^{G} \frac{c_h^{g,\dot{g}} I^{\dot{g}}(t)}{N^{\dot{g}}} \left(1 - e^{-\kappa \frac{n_h^{g,\dot{g}} x_h^{g,\dot{g}}}{1+n_h^{g,\dot{g}}}}\right) \\
+ \mu(1-\alpha^g) \sum_{\dot{g}=1}^{G} \frac{c_h^{g,\dot{g}} I^{\dot{g}}(t)}{N^{\dot{g}}} \left(1 - e^{-\kappa \frac{(1-\alpha^g) n_h^{g,\dot{g}} x_h^{g,\dot{g}}}{1+(1-\alpha^g) n_h^{g,\dot{g}}}}\right) \\
+ \mu \alpha^g \sum_{\dot{g}=1}^{G} \frac{c_w^{g,\dot{g}} I^{\dot{g}}(t)}{N^{\dot{g}}} \left(1 - e^{-\kappa \frac{\alpha^g n_w^{g,\dot{g}} x_w^{g,\dot{g}}}{1+\alpha^g n_w^{g,\dot{g}}}}\right).
\end{aligned} \tag{22}
$$

Equation (22) accounts for infection contributed through interactions occurring with infected individuals within and outside the home by accounting for transmission within and between groups.

We computed the time-dependent reproduction number, $\mathscr{R}_t$, and basic reproduction number, $\mathscr{R}_0$, using the NGM formula for the discrete-time model [4, 5]. For the *SEIR*-type model (17), the infected compartments (E, I) are used to derive the reproduction number. For the NGM, the components of transmissions in the model are given as:

$$
\begin{aligned}
\mathscr{F}_E^g = \mu S^g(t) \sum_{\dot{g}=1}^{G} \Bigg[ \frac{c_h^{g,\dot{g}} I^{\dot{g}}(t)}{N^{\dot{g}}} \left(1 - e^{-\kappa \frac{n_h^{g,\dot{g}} x_h^{g,\dot{g}}}{1+n_h^{g,\dot{g}}}}\right) + (1-\alpha^g) \frac{c_h^{g,\dot{g}} I^{\dot{g}}(t)}{N^{\dot{g}}} \left(1 - e^{-\kappa \frac{(1-\alpha^g) n_h^{g,\dot{g}} x_h^{g,\dot{g}}}{1+(1-\alpha^g) n_h^{g,\dot{g}}}}\right) \\
+ \alpha^g \frac{c_w^{g,\dot{g}} I^{\dot{g}}(t)}{N^{\dot{g}}} \left(1 - e^{-\kappa \frac{\alpha^g n_w^{g,\dot{g}} x_w^{g,\dot{g}}}{1+\alpha^g n_w^{g,\dot{g}}}}\right) \Bigg], \\
\text{for } g, \dot{g} = 1, 2, \ldots, G.
\end{aligned} \tag{23}
$$

The transition components in the model are

$$
\begin{aligned}
\mathscr{T}_{E^g} &= (1-\sigma) E^g(t), \\
\mathscr{T}_{I^g} &= (1-\gamma) I^g(t) + \sigma E^g(t), \\
&\text{for } g, \dot{g} = 1, 2, \ldots, G.
\end{aligned} \tag{24}
$$

The matrices of transmissions, $F$, and transitions, $T$, are given by partial derivatives of $\mathscr{F}$ and $\mathscr{T}$ with respect to $E^1,...,E^G,I^1,...,I^G$.

The matrix of transmission is:

$$
F = \mu \begin{pmatrix}
0 & 0 & \ldots & 0 & F_{11} & F_{12} & \ldots & F_{1G} \\
0 & 0 & \ldots & 0 & F_{21} & F_{22} & \ldots & F_{2G} \\
\vdots & \vdots & \ddots & \vdots & \vdots & \vdots & \ddots & \vdots \\
0 & 0 & \ldots & 0 & F_{G1} & F_{G2} & \ldots & F_{GG} \\
0 & 0 & \ldots & 0 & 0 & 0 & \ldots & 0 \\
0 & 0 & \ldots & 0 & 0 & 0 & \ldots & 0 \\
\vdots & \vdots & \ddots & \vdots & \vdots & \vdots & \ddots & \vdots \\
0 & 0 & \ldots & 0 & 0 & 0 & \ldots & 0
\end{pmatrix}, \tag{25}
$$

with:

$$
F_{ij} = \frac{S^i(t)}{N^j} \left[ c_h^{i,j} \left(1 - e^{-\kappa \frac{n_h^{i,j} x_h^{i,j}}{1+n_h^{i,j}}}\right) + (1-\alpha^i) c_h^{i,j} \left(1 - e^{-\kappa \frac{(1-\alpha^i) n_h^{i,j} x_h^{i,j}}{1+(1-\alpha^i) n_h^{i,j}}}\right) + \alpha^i c_w^{i,j} \left(1 - e^{-\kappa \frac{\alpha^i n_w^{i,j} x_w^{i,j}}{1+\alpha^i n_w^{i,j}}}\right) \right] \tag{26}
$$

The matrix of transition is:

$$T=\begin{pmatrix}1-\sigma & 0 & \dots & 0 & 0 & 0 & \dots & 0\\ 0 & 1-\sigma & \dots & 0 & 0 & 0 & \dots & 0\\ \vdots & \vdots & \ddots & & & \vdots & \vdots & \vdots\\ 0 & 0 & \dots & 1-\sigma & 0 & 0 & \dots & 0\\ \sigma & 0 & \dots & 0 & 1-\gamma & 0 & \dots & 0\\ 0 & \sigma & \dots & 0 & 0 & 1-\gamma & \dots & 0\\ \vdots & \vdots & \ddots & & & \vdots & \ddots & \vdots\\ 0 & 0 & \dots & \sigma & 0 & 0 & \dots & 1-\gamma\end{pmatrix}. \tag{27}$$

From the diagonal elements of $T$, the condition $\rho(T) < 1$ for the discrete-time model [4, 5] is satisfied. Furthermore, we compute $\mathbb{I}-T$ as

$$\mathbb{I}-T=\begin{pmatrix}\sigma & 0 & \dots & 0 & 0 & 0 & \dots & 0\\ 0 & \sigma & \dots & 0 & 0 & 0 & \dots & 0\\ \vdots & \vdots & \ddots & & & \vdots & \vdots & \vdots\\ 0 & 0 & \dots & \sigma & 0 & 0 & \dots & 0\\ -\sigma & 0 & \dots & 0 & \gamma & 0 & \dots & 0\\ 0 & -\sigma & \dots & 0 & 0 & \gamma & \dots & 0\\ \vdots & \vdots & \ddots & & & \vdots & \ddots & \vdots\\ 0 & 0 & \dots & -\sigma & 0 & 0 & \dots & \gamma\end{pmatrix} \tag{28}$$

To find the inverse of the matrix $\mathbb{I}-T$, we use the block matrix inversion formula for a partitioned matrix. The matrix can be partitioned into four blocks as follows:

$$\mathbb{I}-T=\begin{pmatrix}A & B\\ C & D\end{pmatrix}. \tag{29}$$

If $A$ and $D$ are invertible matrices, then the formula for the inverse of the partitioned matrix [6] is given by:

$$(\mathbb{I}-T)^{-1}=\begin{pmatrix}(A-BD^{-1}C)^{-1} & -A^{-1}BD^{-1}\\ -D^{-1}C(A-BD^{-1}C)^{-1} & D^{-1}+D^{-1}CA^{-1}BD^{-1}\end{pmatrix}, \tag{30}$$

where in this model, $A$, $B$, $C$, and $D$ are defined as follows:

$$A=\begin{pmatrix}\sigma & 0 & \dots & 0\\ 0 & \sigma & \dots & 0\\ \vdots & \vdots & \ddots & \vdots\\ 0 & 0 & \dots & \sigma\end{pmatrix},\quad B=0_{G\times G},\quad C=\begin{pmatrix}-\sigma & 0 & \dots & 0\\ 0 & -\sigma & \dots & 0\\ \vdots & \vdots & \ddots & \vdots\\ 0 & 0 & \dots & -\sigma\end{pmatrix},\quad D=\begin{pmatrix}\gamma & 0 & \dots & 0\\ 0 & \gamma & \dots & 0\\ \vdots & \vdots & \ddots & \vdots\\ 0 & 0 & \dots & \gamma\end{pmatrix}. \tag{31}$$

Since the upper block $B$ is a square matrix of zeros, the block matrix inversion formula becomes:

$$(\mathbb{I}-T)^{-1}=\begin{pmatrix}A^{-1} & 0\\ -D^{-1}CA^{-1} & D^{-1}\end{pmatrix}. \tag{32}$$

1. **Inverse of $A$:**

$$A^{-1}=\begin{pmatrix}\frac{1}{\sigma} & 0 & \dots & 0\\ 0 & \frac{1}{\sigma} & \dots & 0\\ \vdots & \vdots & \ddots & \vdots\\ 0 & 0 & \dots & \frac{1}{\sigma}\end{pmatrix} \tag{33}$$

2. **Inverse of $D$:**

$$D^{-1} = \begin{pmatrix} \frac{1}{\gamma} & 0 & \dots & 0 \\ 0 & \frac{1}{\gamma} & \dots & 0 \\ \vdots & \vdots & \ddots & \vdots \\ 0 & 0 & \dots & \frac{1}{\gamma} \end{pmatrix} \tag{34}$$

3. **Compute $-D^{-1}CA^{-1}$:**

$$-D^{-1}CA^{-1} = -\begin{pmatrix} \frac{1}{\gamma} & 0 & \dots & 0 \\ 0 & \frac{1}{\gamma} & \dots & 0 \\ \vdots & \vdots & \ddots & \vdots \\ 0 & 0 & \dots & \frac{1}{\gamma} \end{pmatrix} \begin{pmatrix} -\sigma & 0 & \dots & 0 \\ 0 & -\sigma & \dots & 0 \\ \vdots & \vdots & \ddots & \vdots \\ 0 & 0 & \dots & -\sigma \end{pmatrix} \begin{pmatrix} \frac{1}{\sigma} & 0 & \dots & 0 \\ 0 & \frac{1}{\sigma} & \dots & 0 \\ \vdots & \vdots & \ddots & \vdots \\ 0 & 0 & \dots & \frac{1}{\sigma} \end{pmatrix} \tag{35}$$

4. **Combine the results:**

$$(\mathbb{I}-T)^{-1} = \begin{pmatrix} \frac{1}{\sigma} & 0 & \dots & 0 & 0 & 0 & \dots & 0 \\ 0 & \frac{1}{\sigma} & \dots & 0 & 0 & 0 & \dots & 0 \\ \vdots & \vdots & \ddots & & & \vdots & \vdots & \vdots \\ 0 & 0 & \dots & \frac{1}{\sigma} & 0 & 0 & \dots & 0 \\ \frac{1}{\gamma} & 0 & \dots & 0 & \frac{1}{\gamma} & 0 & \dots & 0 \\ 0 & \frac{1}{\gamma} & \dots & 0 & 0 & \frac{1}{\gamma} & \dots & 0 \\ \vdots & \vdots & \ddots & & & \vdots & \ddots & \vdots \\ 0 & 0 & \dots & \frac{1}{\gamma} & 0 & 0 & \dots & \frac{1}{\gamma} \end{pmatrix} \tag{36}$$

To compute the NGM, let

$$F = \begin{pmatrix} F11 & F12 \\ F21 & F22 \end{pmatrix}, \quad V = (\mathbb{I}-T)^{-1} = \begin{pmatrix} V11 & V12 \\ V21 & V22 \end{pmatrix} \tag{37}$$

Where

$$F11 = 0_{G\times G}, \quad F12 = \mu \begin{pmatrix} F_{11} & F_{12} & \dots & F_{1G} \\ F_{21} & F_{22} & \dots & F_{2G} \\ \vdots & \vdots & \ddots & \vdots \\ F_{G1} & F_{G2} & \dots & F_{GG} \end{pmatrix}, \tag{38}$$

$$F21 = 0_{G\times G}, \quad F22 = 0_{G\times G} \tag{39}$$

$$V11 = \begin{pmatrix} \frac{1}{\sigma} & 0 & \dots & 0 \\ 0 & \frac{1}{\sigma} & \dots & 0 \\ \vdots & \vdots & \ddots & \vdots \\ 0 & 0 & \dots & \frac{1}{\sigma} \end{pmatrix}, \quad V12 = 0_{G\times G} \tag{40}$$

$$V21 = \begin{pmatrix} \frac{1}{\gamma} & 0 & \dots & 0 \\ 0 & \frac{1}{\gamma} & \dots & 0 \\ \vdots & \vdots & \ddots & \vdots \\ 0 & 0 & \dots & \frac{1}{\gamma} \end{pmatrix}, \quad V22 = \begin{pmatrix} \frac{1}{\gamma} & 0 & \dots & 0 \\ 0 & \frac{1}{\gamma} & \dots & 0 \\ \vdots & \vdots & \ddots & \vdots \\ 0 & 0 & \dots & \frac{1}{\gamma} \end{pmatrix} \tag{41}$$

The NGM is therefore given by

$$F(\mathbb{I}-T)^{-1} = FV = \begin{pmatrix} F11 & F12 \\ F21 & F22 \end{pmatrix} \begin{pmatrix} V11 & V12 \\ V21 & V22 \end{pmatrix} = \begin{pmatrix} F11V11+F12V21 & F11V12+F12V22 \\ F21V11+F22V21 & F21V12+F22V22 \end{pmatrix} \tag{42}$$

$$F(\mathbb{I}-T)^{-1} = \frac{\mu}{\gamma} \begin{pmatrix} F_{11} & F_{12} & \dots & F_{1G} & F_{11} & F_{12} & \dots & F_{1G} \\ F_{21} & F_{22} & \dots & F_{2G} & F_{21} & F_{22} & \dots & F_{2G} \\ \vdots & \vdots & \ddots & & & \vdots & \vdots & \vdots \\ F_{G1} & F_{G2} & \dots & F_{GG} & F_{G1} & F_{G2} & \dots & F_{GG} \\ 0 & 0 & \dots & 0 & 0 & 0 & \dots & 0 \\ 0 & 0 & \dots & 0 & 0 & 0 & \dots & 0 \\ \vdots & \vdots & & & & \vdots & \ddots & \vdots \\ 0 & 0 & \dots & 0 & 0 & 0 & \dots & 0 \end{pmatrix} \tag{43}$$

The overall basic reproduction number $\mathscr{R}_0$ is the spectral radius of the next generation matrix (43). Let the NGM (43) be denoted by $\mathbb{A}$, so that the matrix is written as:

$$\mathbb{A} = \begin{pmatrix} A & A \\ 0 & 0 \end{pmatrix}$$

Where

$$A = \frac{\mu}{\gamma} \begin{pmatrix} F_{11} & F_{12} & \dots & F_{1G} \\ F_{21} & F_{22} & \dots & F_{2G} \\ \vdots & \vdots & \ddots & \vdots \\ F_{G1} & F_{G2} & \dots & F_{GG} \end{pmatrix} \tag{44}$$

The elements 0s are block matrices of the size of $A$.

To find the eigenvalues of the matrix $\mathbb{A}$, we need to solve the characteristic equation, which is given by $\det(\mathbb{A}-\lambda\mathbb{I}) = 0$.

The matrix is given as

$$\mathbb{A}-\lambda\mathbb{I} = \begin{pmatrix} A-\lambda I & A \\ 0 & -\lambda I \end{pmatrix}$$

Here, $I$ represents the identity matrix of the size of $A$, while $\lambda$ are the eigenvalues. Now, the determinant is given by:

$$\det(\mathbb{A}-\lambda\mathbb{I}) = \det \begin{pmatrix} A-\lambda I & A \\ 0 & -\lambda I \end{pmatrix}$$

$$= \det(A-\lambda I)\cdot\det(-\lambda I)$$

$$= \det(A-\lambda I)\cdot(-\lambda)^{\text{size of } A}$$

Setting this determinant equal to zero and solving for $\lambda$ gives

$$\det(A-\lambda I)\cdot(-\lambda)^{\text{size of } A} = 0$$

This equation has eigenvalues corresponding to the solutions for $\lambda$. Note that the eigenvalues of $A-\lambda I$ are found by setting the determinant of $A-\lambda I$ equal to zero. The $(-\lambda)^{\text{size of } A}$ part corresponds to the eigenvalues associated with the zero block.

The reproduction number is given by the solutions to $\det(A-\lambda I) = 0$. However, the analytical expression of this term is difficult to derive, especially when $G > 2$, as such numerical computation is preferable to determine the value of the reproduction number.

The time-dependent reproduction number, defined as the average number of secondary infections created in the population (regardless of the demographic group of the infected and the source), is the spectral radius of the matrix (44) computed as:

$$\mathscr{R}_t = \rho(A). \tag{45}$$

By evaluating at the disease-free equilibrium (taking the susceptible populations across all groups to be approximately the total population sizes), the basic reproduction number is given as:

$$\mathscr{R}_0 = \rho(A)_{S^g \approx N^g} \forall g. \tag{46}$$

#### 1.4 Source-to-Sink reproduction number

One of the most crucial motivations for developing mathematical models in epidemiology is to understand the mechanisms needed to control or prevent outbreaks that threaten lives and economic development. Although $\mathscr{R}_0$ can be useful when designing control strategies during outbreaks, the overall $\mathscr{R}_0$ does not give insights into the individual contributions of each group in the spread of pathogens [7].

We define the following:

- The time-dependent *source to sink* reproduction number of group $\dot{g}$ (source) on group $g$ (sink) is the $F_{i,j}$ element of the matrix (44) (where $g$ corresponds to $i$ and $\dot{g}$ corresponds to $j$) computed as:

$$\overset{\dot{g}\to g}{\mathscr{R}_t} = \frac{\mu S^g(t)}{\gamma N^{\dot{g}}}\left[c_h^{g,\dot{g}}\left(1-e^{-\kappa\frac{n_h^{g,\dot{g}}x_h^{g,\dot{g}}}{1+n_h^{g,\dot{g}}}}\right)+(1-\alpha^g)c_h^{g,\dot{g}}\left(1-e^{-\kappa\frac{(1-\alpha^g)n_h^{g,\dot{g}}x_h^{g,\dot{g}}}{1+(1-\alpha^g)n_h^{g,\dot{g}}}}\right)+\alpha^g c_w^{g,\dot{g}}\left(1-e^{-\kappa\frac{\alpha^g n_w^{g,\dot{g}}x_w^{g,\dot{g}}}{1+\alpha^g n_w^{g,\dot{g}}}}\right)\right]. \tag{47}$$

- The time-dependent *source* reproduction number of group $\dot{g}$ (source) on all other groups is the sum of the $\dot{g}^{th}$ column of the matrix (44) computed as:

$$\overset{\dot{g}\to All}{\mathscr{R}_t} = \sum_g \frac{\mu S^g(t)}{\gamma N^{\dot{g}}}\left[c_h^{g,\dot{g}}\left(1-e^{-\kappa\frac{n_h^{g,\dot{g}}x_h^{g,\dot{g}}}{1+n_h^{g,\dot{g}}}}\right)+(1-\alpha^g)c_h^{g,\dot{g}}\left(1-e^{-\kappa\frac{(1-\alpha^g)n_h^{g,\dot{g}}x_h^{g,\dot{g}}}{1+(1-\alpha^g)n_h^{g,\dot{g}}}}\right)+\alpha^g c_w^{g,\dot{g}}\left(1-e^{-\kappa\frac{\alpha^g n_w^{g,\dot{g}}x_w^{g,\dot{g}}}{1+\alpha^g n_w^{g,\dot{g}}}}\right)\right]. \tag{48}$$

- The time-dependent *sink* reproduction number of group $g$ (sink) is the sum of the $g^{th}$ row of the matrix (44) computed as:

$$ {}^{All\to g}\mathscr{R}_t = \sum_{\hat{g}} \frac{\mu S^g(t)}{\gamma N^{\hat{g}}} \left[ c_h^{g,\hat{g}} \left( 1 - e^{-\kappa \frac{n_h^{g,\hat{g}} x_h^{g,\hat{g}}}{1+n_h^{g,\hat{g}}}} \right) + (1-\alpha^g) c_h^{g,\hat{g}} \left( 1 - e^{-\kappa \frac{(1-\alpha^g) n_h^{g,\hat{g}} x_h^{g,\hat{g}}}{1+(1-\alpha^g) n_h^{g,\hat{g}}}} \right) + \alpha^g c_w^{g,\hat{g}} \left( 1 - e^{-\kappa \frac{\alpha^g n_w^{g,\hat{g}} x_w^{g,\hat{g}}}{1+\alpha^g n_w^{g,\hat{g}}}} \right) \right]. \tag{49} $$

The reproduction numbers derived using this framework rely primarily on two fundamental epidemic parameters: the transmission probability, $\mu$, and the recovery probability, $\gamma$. However, this framework reveals how social and demographic parameters impact its value. These reproduction numbers could serve as important indicators for public health officials and policymakers in formulating effective targeted strategies for pathogen control and containment.

From the analytical expressions of these quantities, the *source* and *sink* reproduction numbers give information that the basic reproduction number may not explicitly give, as they reveal the contribution to and from each group in the pathogen propagation. Also, the *source-to-sink* reproduction number gives additional information that is not readily available using the basic, the *source*, and the *sink* reproduction numbers, as it gives the information about the impact of each group on the other groups in the spread of pathogens.

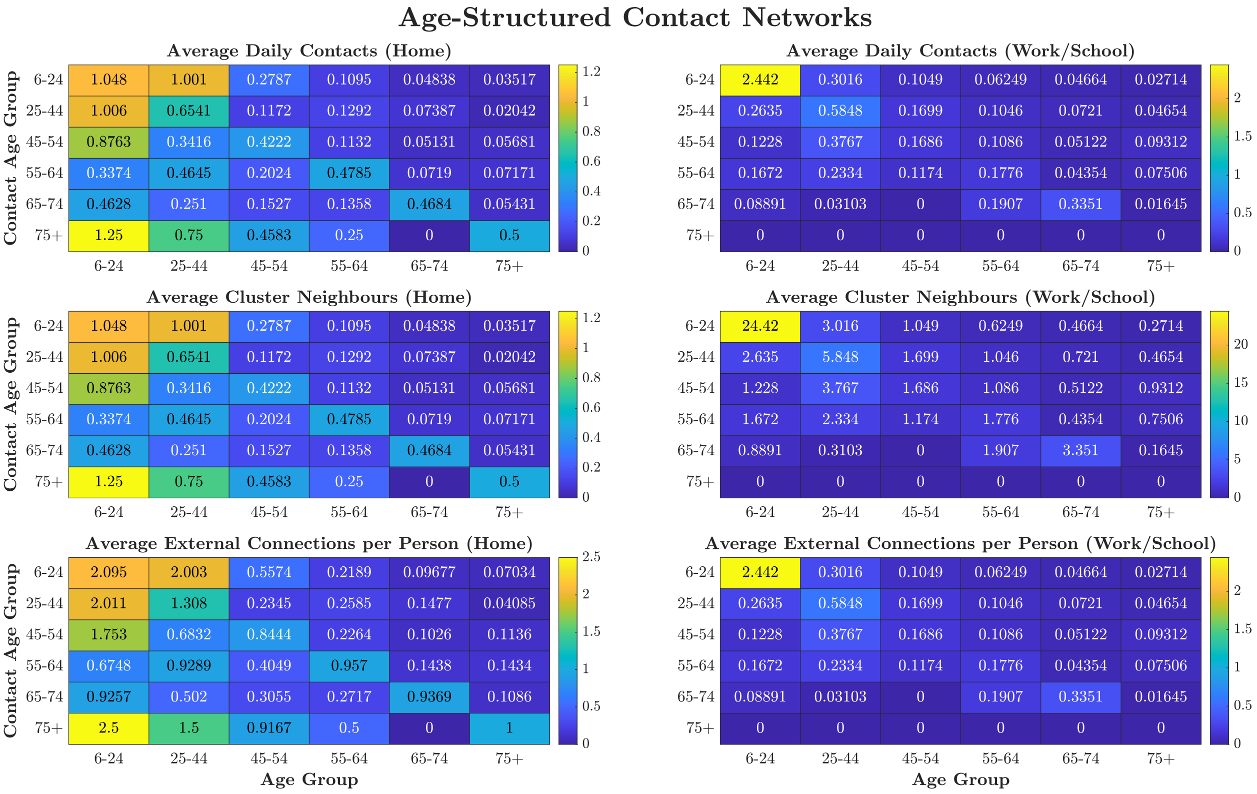


**Figure 2.** Age-structured contact networks derived from UK social mixing data illustrating average daily contacts, cluster neighbours, and external connections for home and work/school environments.

**Table 1.** Model parameters. (Notes: $\sigma$ and $\gamma$ shown are probabilities per time step obtained from rates via $P_r = 1 - e^{-rt}$.)

| Parameter | Description | Value |
|---|---|---|
| $\alpha$ | mobility probability | 0.72 |
| $\sigma$ | progression probability | 0.1813 |
| $\gamma$ | recovery probability | 0.3935 |
| $\kappa$ | coupling constant | 0.6031 |

**Table 2.** Infection Hospitalisation Risk (IHR) by Age - COVID-19

| Age Groups | IHR % [9] |
|---|---|
| 6–24 | 0.0036 |
| 25–44 | 0.0139 |
| 45–54 | 0.0218 |
| 55–64 | 0.0434 |
| 65–74 | 0.0997 |
| $\geq 75$ | 0.3393 |

To analyse the effect of work/school closure for different social connectivities, we changed the baseline household and work/school external connections $(x_h, x_w)$ along with the mobility probability $(\alpha)$. The focus is to see whether or not increasing mobility to work/school would always increase the epidemic final sizes, particularly considering that different populations may have different connectivity structures. We used **Algorithm** 1 to compute the final sizes for each combination of modified parameters $\alpha$ and $x_h/x_w$, using different percentages (0–100%) of the baseline values.

**Algorithm 1** Computing Epidemic Final Size for Varying Mobility $(\alpha)$ and Household/Work External $(x_h/x_w)$Connections

**Input:**
Model parameters
Function for running SEIR Model
**Output:**
3D array `FinalSize` containing epidemic final sizes for each combination of scaled $\alpha$, $x_h/x_w$, and age group.
**Initialize:**
Generate `scale_factors` as 15 evenly spaced points from 0 to 1 (representing 0% to 100% of baseline values). *The choice of 15 points is for smoothness*
Construct 2D grids `Alpha` and `Xh/Xw` using `meshgrid(scale_factors, scale_factors)` to represent all combinations of scaled $\alpha$ and $x_h/x_w$.
Initialize `FinalSize` as a zero array of size (`size(Alpha, 1)`, `size(Alpha, 2)`, `Number of age groups`) to store final sizes for each parameter combination and age group.
**Computation:**
**for** each index $i$ from 1 to `size(Alpha, 1)` **do** ▷ Loop over rows of `Alpha` (scaled $\alpha$ values)
  **for** each index $j$ from 1 to `size(Xh/Xw, 2)` **do** ▷ Loop over columns of `Xh/Xw` (scaled $x_h/x_w$ values)
    Set $\alpha_{new} \leftarrow$ `Alpha(i, j)` $\times\ \alpha$. ▷ Scale $\alpha$ to 0–100% of baseline
    Set $x_{hnew}/x_{wnew} \leftarrow$ `Xh/Xw(i, j)` $\times\ x_h/x_w$. ▷ Scale $x_h$ to 0–100% of baseline
    Compute model output: Call simulation function. ▷ Run epidemiological model with the new parameter set
    Store `FinalSize(i, j, :)` $\leftarrow$ `Total recovered.` ▷ Save final recovered individuals for all age groups as the final size
  **end for**
**end for**
**Return:** `FinalSize`.

### Acknowledgements

### Acknowledgements

This research is fully funded by the Institute for Global Pandemic Planning, Warwick Medical School, University of Warwick UK.

### Author contributions statement

Conceptualization: M. L. S.
Methodology: M. L. S.
Supervision: A.S. & K. S. R.
Software: K. S. R. & M. L. S.
Visualisation: K S. R & M. L. S.
Writing – original draft: M. L. S.
Writing – review & editing: A. S., K. S. R. & M. L. S.

### Data availability

The MATLAB code used to generate the figures is available on Zenodo `https://doi.org/10.5281/zenodo.17605791`

### Ethics declarations

This study focuses on the theoretical development of a mathematical model for the spread of a hypothetical infectious disease. No data was collected and analysed.

### Additional information

**Competing interests**. The authors declare no competing interests.
